# Drug-induced glucose metabolism disorders: A disproportionality analysis based on the FAERS database

**DOI:** 10.1101/2025.08.26.25334521

**Authors:** Zhongxiang Zhang, Qiaoying Li, Tao Huang, Xuping Yang, Xinyun Du, Xinyi Deng, Shurong Wang, Jie Zhou

## Abstract

**Background:** Glucose metabolism disorders encompass abnormalities in glucose digestion, absorption, transport, utilization, and regulation, leading to broad physiological and pathological consequences. Although drug-induced disturbances are increasingly documented, they remain under-recognized in clinical practice and drug labeling.

**Methods:** This disproportionality analysis used publicly available data from the FDA Adverse Event Reporting System (FAERS), covering reports from Q4 2004 to Q1 2025. After data cleaning and standardization, four disproportionality methods (ROR, PRR, MGPS, BCPNN) were applied to detect signals. A signal was considered positive only if all method thresholds were met (ROR: n ≥ 3, lower 95% CI > 1; PRR: χ² ≥ 4, lower 95% CI > 1; MGPS: EBGM05 > 2; BCPNN: IC025 > 0). A descriptive analysis of clinical characteristics and a case-by-case assessment were also performed.

**Results:** Among 22,775,812 reports, 204,236 were related to glucose metabolism disorders and involved 1,827 drugs. A total of 128 drugs showed positive signals. The most frequent classes were anti-diabetic drugs (38%), antineoplastic agents (9.3%), renin-angiotensin system drugs (8.6%), and systemic corticosteroids (4.7%). Notably, several drugs, including basiliximab, enfortumab vedotin, and mercaptopurine, lack explicit warnings regarding glucose metabolism disorders.

**Conclusion:** This study identifies potential safety signals that require further clinical validation. These findings emphasize the need for improved monitoring and timely updates to drug labeling, particularly for high-risk populations. Disproportionality analysis is hypothesis-generating and should be interpreted with caution.

## 1. Introduction

Glucose metabolism is a fundamental physiological process that underpins cellular energy homeostasis and various key biological functions, including cell proliferation, signal transduction, redox balance, and the biosynthesis of biomacromolecules(^1, 2^). Its proper functioning relies on the precise regulation of blood glucose levels, primarily mediated by insulin and glucagon. Disruption of this regulatory network can lead to disorders of glucose metabolism, which encompass a range of abnormalities from hypoglycemia to hyperglycemia. Common conditions include chronic hyperglycemia, such as Type 2 Diabetes Mellitus, prediabetes, and gestational diabetes mellitus, as well as hypoglycemia resulting from insulin excess, inadequate glucagon, or defective glucose-raising mechanisms(^3^). These disorders constitute a significant and growing global health burden(4).

Many drugs have been shown to interfere with glucose homeostasis, leading to the induction or exacerbation of glucose metabolism disorders, commonly referred to as drug-induced glucose metabolism disorders. These disorders manifest as hyperglycemia (including drug-induced diabetes) or hypoglycemia(5–7). The effects and severity of drug-induced disturbances depend on a combination of pharmacologic properties, including mechanism of action, dosage, and regimen, as well as individual patient risk factors such as baseline glucose levels, obesity, genetic predisposition, and renal function(8, 9). For example, glucocorticoid medications are a well-known cause of pharmacological hyperglycemia, primarily by inducing insulin resistance and promoting hepatic gluconeogenesis(10). In addition, certain antipsychotics, such as olanzapine and quetiapine, are associated with weight gain, insulin resistance, and an increased risk of new-onset diabetes(11). Furthermore, inappropriate use of insulin or insulinotropic agents, such as sulfonylureas, significantly contributes to iatrogenic hypoglycemia(12) Additionally, some antimicrobials, including fluoroquinolones, and antiarrhythmics, such as propranolol, have also been linked to hypoglycemic events(13, 14). Given these risks, the identification and effective management of drug-induced glucose metabolism disorders have become critical areas of clinical practice that require heightened vigilance. This is especially true for high-risk populations, such as the elderly or patients with pre-existing metabolic syndrome or diabetes(4, 15).

While certain drugs, such as glucocorticoids and specific antipsychotics, are strongly associated with inducing glucose metabolism disorders, many other drugs with potential disruptive effects remain underrecognized or unquantified. This study aimed to fill this knowledge gap by systematically identifying drugs associated with glucose metabolism disorders using real-world pharmacovigilance data. Pharmacovigilance data provide a more accurate reflection of real-world drug use compared to laboratory or clinical trial data, making them indispensable for post-marketing surveillance(16). Databases like the FDA Adverse Event Reporting System (FAERS), which aggregate spontaneous reports of adverse drug events, are particularly valuable for detecting potential safety signals. Their significance lies in their large volume, diverse information, and public accessibility(^17^). We applied disproportionality analysis to the entire FAERS database as the background reference population to identify significant reporting associations. Utilizing disproportionality analysis on such databases is a well-established method for systematically mining data to identify novel or underappreciated statistical associations between drugs and adverse events(^17^). This approach is particularly well-suited to bridge the knowledge gap concerning the broader landscape of drugs that may induce glucose metabolism disorders, extending beyond those that are already well-documented.

## 2. Materials and methods

### 2.1 Data source and study design

We conducted a retrospective observational study using a disproportionality analysis design on data from the publicly available FAERS database. The overall study design included a primary disproportionality analysis and supplementary descriptive analyses of patient characteristics and time-to-onset. A case-by-case review was not performed due to the large number of signals identified; instead, we focused on a detailed descriptive analysis and contextualization with existing literature. Data were extracted on July 10, 2025, covering reports from the fourth quarter of 2004 to the first quarter of 2025. The FAERS database is maintained by the U.S. Food and Drug Administration and contains spontaneous reports from healthcare professionals, consumers, and manufacturers worldwide, though the majority originate from the United States. Adverse events and medication errors in the FAERS database were coded using terms from the Medical Dictionary for Regulatory Activities (MedDRA). In this study, the most recent version of the MedDRA dictionary (MedDRA 27.1) was employed to code the names of adverse events recorded in the FAERS database.

This study conducted a retrospective observational analysis utilizing the public database of the US FDA Adverse Event Reporting System. All data within this database have undergone rigorous anonymization processes and do not contain any personally identifiable information, making it a publicly accessible scientific resource. In accordance with the exemption clause for the secondary use of public databases as outlined in international ethical guidelines, this study did not involve individual intervention or harm; therefore, it did not require review by an ethics review committee.

### 2.2 Data cleansing

Duplicate, withdrawn, and deleted reports may arise due to the spontaneous submission of data to the database. Consequently, the FDA’s official guidance document delineates specific rules for identifying and removing duplicate data and provides a comprehensive list of reports that should be deleted. In this study, we strictly followed the FDA guidance document during the data cleaning process. Specifically, duplicate reports were identified and removed based on the recommended primary key (CASEID and ISR number combinations) and the most recent CASEVERSION was retained for each unique case. The number of reports at each stage of data cleaning is detailed in the Results section (Fig 1).

**Fig 1.**
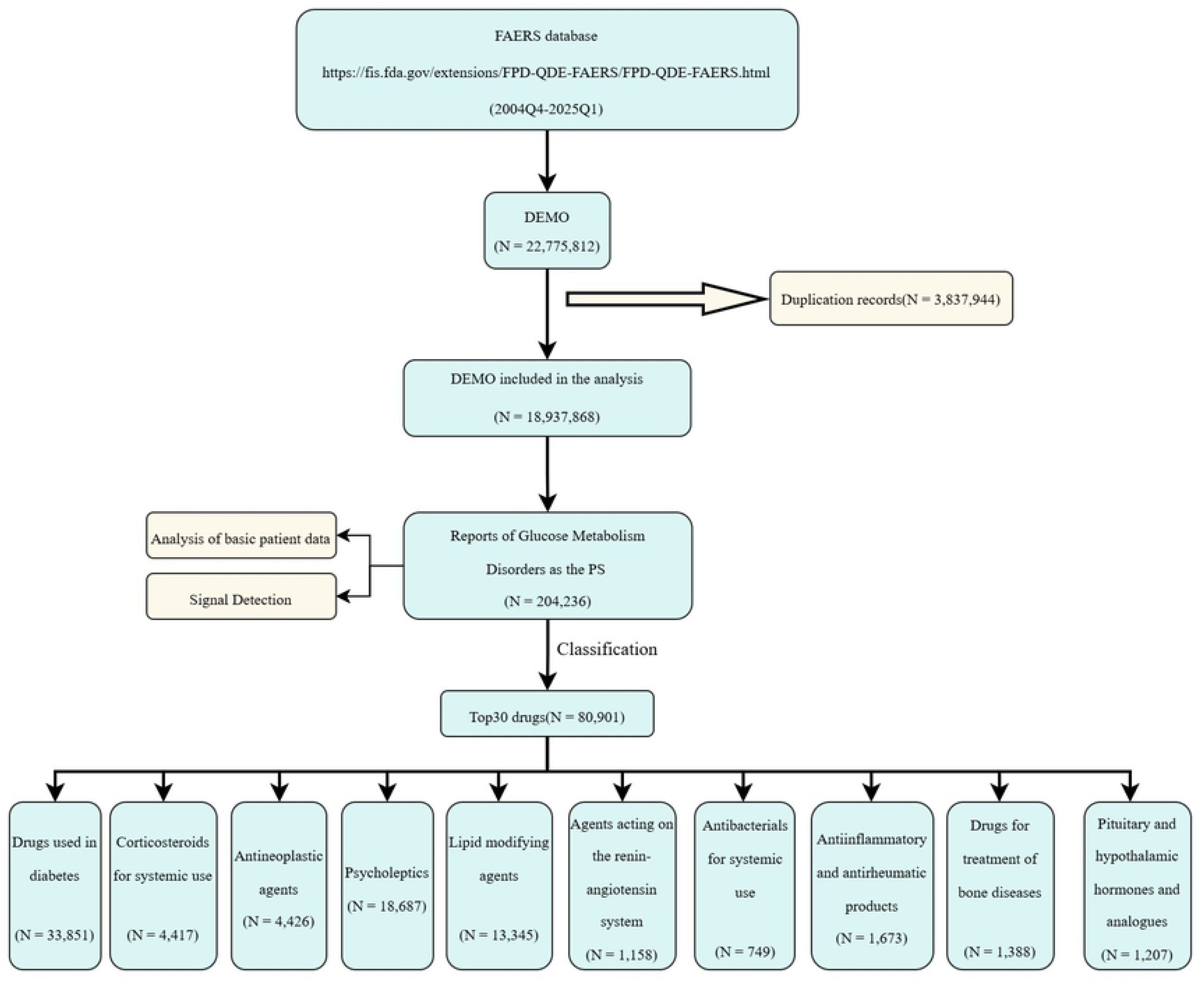
Flow chart for identifying reports of suspected Glucose Me.

### 2.3 Screening the Target Drug Population

Each patient report in the database identifies a unique primary suspected drug (PS). In line with READUS-PV terminology, a ‘drug’ in this analysis refers specifically to the ‘Primary Suspected Drug’, defined as the substance considered by the reporter to be most likely causally associated with the adverse event. Only the primary suspected drug is considered when determining the target drug use population. If a patient’s primary suspected drug corresponds with the target drug under investigation in the background database, the patient is included in the target drug population; otherwise, they are classified into the other drug population. All drug names in the database have been standardized according to the World Health Organization Drug Dictionary and verified using standardized common names.

### 2.4 Statistical analysis

Disproportionality analysis, a widely accepted method for detecting safety signals from individual case safety reports, was a critical component of our methodology. The specific analytical methods were employed in this study included the Reporting Odds Ratio (ROR), Proportional Reporting Ratio (PRR), Multi-item Gamma Poisson Shrinker (MGPS), and Bayesian Confidence Propagation Neural Network (BCPNN), as detailed in S1 Table. We employed these four complementary methods to enhance the robustness of signal detection, as each has different strengths in handling variability and minimizing the risk of false positives. Positive adverse event (AE) signals were identified when the thresholds for all four methods were satisfied: ROR (number of reports [n] ≥ 3, lower 95% confidence interval [CI] > 1); PRR (chi-square [χ²] ≥ 4, lower 95% CI > 1); MGPS [Empirical Bayes Geometric Mean 05 (EBGM05, representing the lower 95% CI) > 2] and BCPNN [Information Component 025 (IC025, representing the lower 95% CI) > 0]. The entire FAERS database during the study period served as the background reference population for calculating expected reporting rates. No sensitivity analyses (e.g., stratification by age, sex, or indication) were performed due to the high proportion of missing data in these fields.

Based on MedDRA terminology, this study conducted disproportionality analyses of adverse events reported in the FAERS at the Preferred Term (PT) level. Utilizing the latest MedDRA version 27.1, the study examined ‘glucose metabolism disorders (MedDRA 10018424) at the High-Level Term (HLT) level and identified 45 related PTs, including type 2 diabetes mellitus, type 1 diabetes mellitus, metabolic syndrome, steroid diabetes, and hypoglycemia (S2 Table).

### 2.5 Statistical analysis software

This study utilized SAS 9.4 for statistical analysis, a software endorsed by the FDA for mining the FAERS database. Data analysis was further facilitated by Navicat (version 16), Microsoft Excel (version 2021), and SPSS (version 27.0.1). The visualization of results was performed using GraphPad (version 9.5). The SAS code used for data cleaning and disproportionality analysis is available from the corresponding author upon reasonable request.

## 3. Results

### 3.1 Descriptive analysis and Participant Flow

#### 3.1.1 The basic process for retrieving adverse event reports of target drugs

A flowchart detailing the number of reports at each stage of data cleaning and inclusion is provided in Fig 1. A total of 22,775,812 reports were retrieved from the FAERS database. After data cleaning and analysis, 204,236 reports related to glucose metabolism disorders were collected. Among these, 1,827 drugs were found to be associated with such disorders. A comprehensive list of the top 30 drugs that met the criteria of all four algorithms (ROR, PRR, MGPS, and BCPNN) was compiled and is presented in order of magnitude in Fig 1. The number of drugs identified by each algorithm is illustrated in Fig 2. Detailed information regarding all 128 drugs that satisfied the criteria of the four algorithms, including the point estimates and 95% confidence intervals for ROR, PRR, EBGM, and IC, is provided in S3 Table.

**Fig 2.**
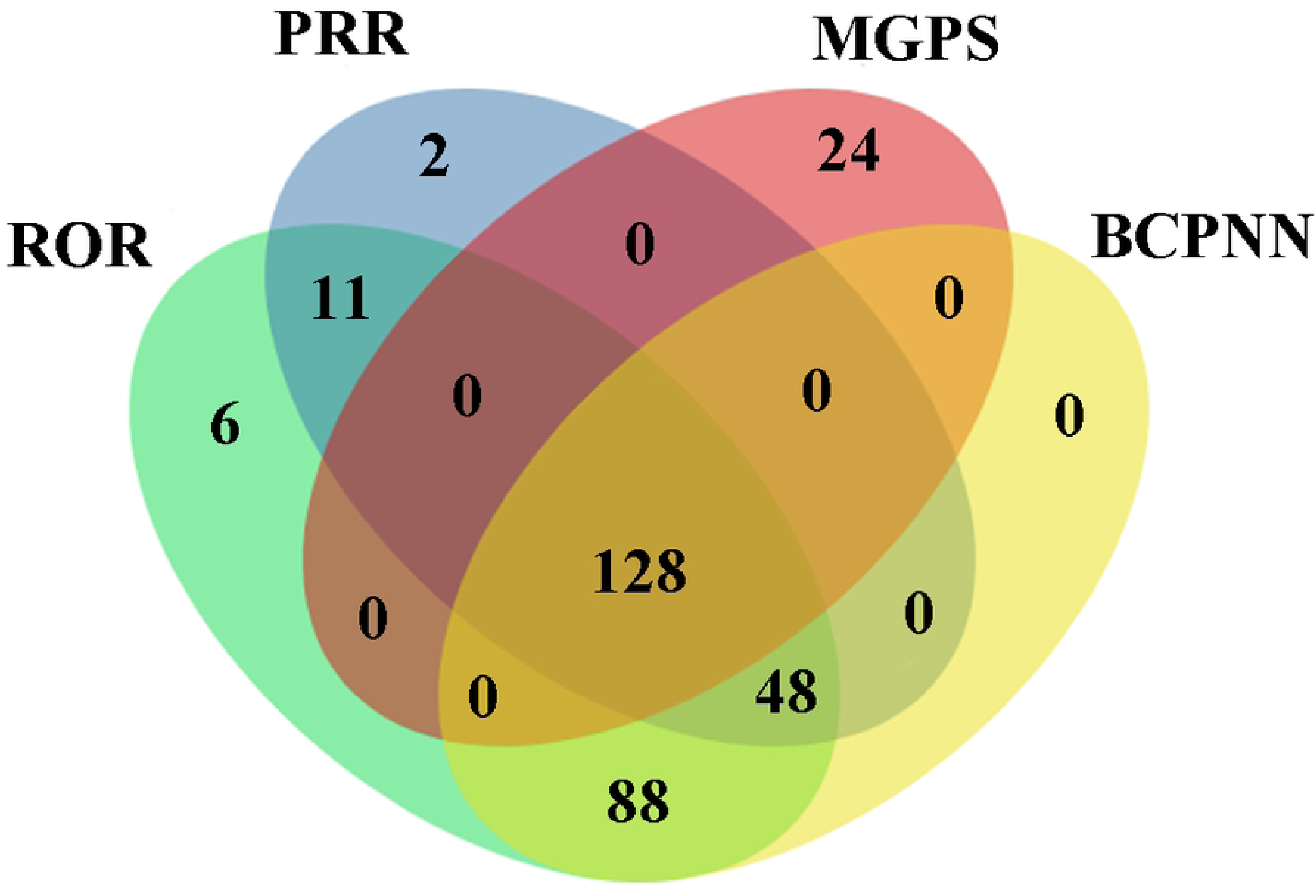
Number of drugs screened by four methods.

#### 3.1.2 Analysis of basic patient data

The analyses were conducted on patients who experienced target adverse events, which were the primary focus of the induction therapy. These adverse events were statistically summarized from the patients’ perspectives. Notably, patients with multiple concurrent adverse events were counted as a single case. The analysis encompassed various indicators, including sex, age, weight, reporter type, reporting country, and clinical outcome. Due to the spontaneous nature of reporting, data were missing for many variables (e.g., weight was missing in 67.1% of reports), limiting the completeness of the descriptive analysis. The demographic and clinical characteristics of the study population were generally consistent with those of the overall FAERS database, supporting the use of the entire FAERS as a valid reference group for disproportionality analysis.

Detailed information regarding patient adverse event reports is presented in Table 1. With respect to sex, females reported 104,292 cases (51.1%) compared to 80,223 cases (39.2%) reported by males, indicating a slightly higher incidence of adverse event reporting among females. This distribution is consistent with the overall FAERS database. In terms of age distribution, the largest proportion of reports came from patients aged 18 to 65 years, accounting for 79,253 reports (38.8%). Regarding weight distribution, 5,568 reports (2.7%) involved patients weighing less than 50 kg, 47,670 reports (23.3%) were from individuals weighing between 50 and 100 kg, and 13,861 reports (6.8%) were from patients exceeding 100 kg. The data for this study were submitted by multiple countries, with the United States contributing the highest number of reports (106,294, 52.1%), followed by Canada (14,478, 7.1%) and Japan (11,485, 5.6%). By indication, diabetes-related diseases accounted for the highest proportion with 25,094 reports (12.2%). In terms of outcomes, “Other” outcomes were the most frequently reported (111,310, 54.5%), followed by “Hospitalization” (62,512, 30.6%).

**Table 1.**
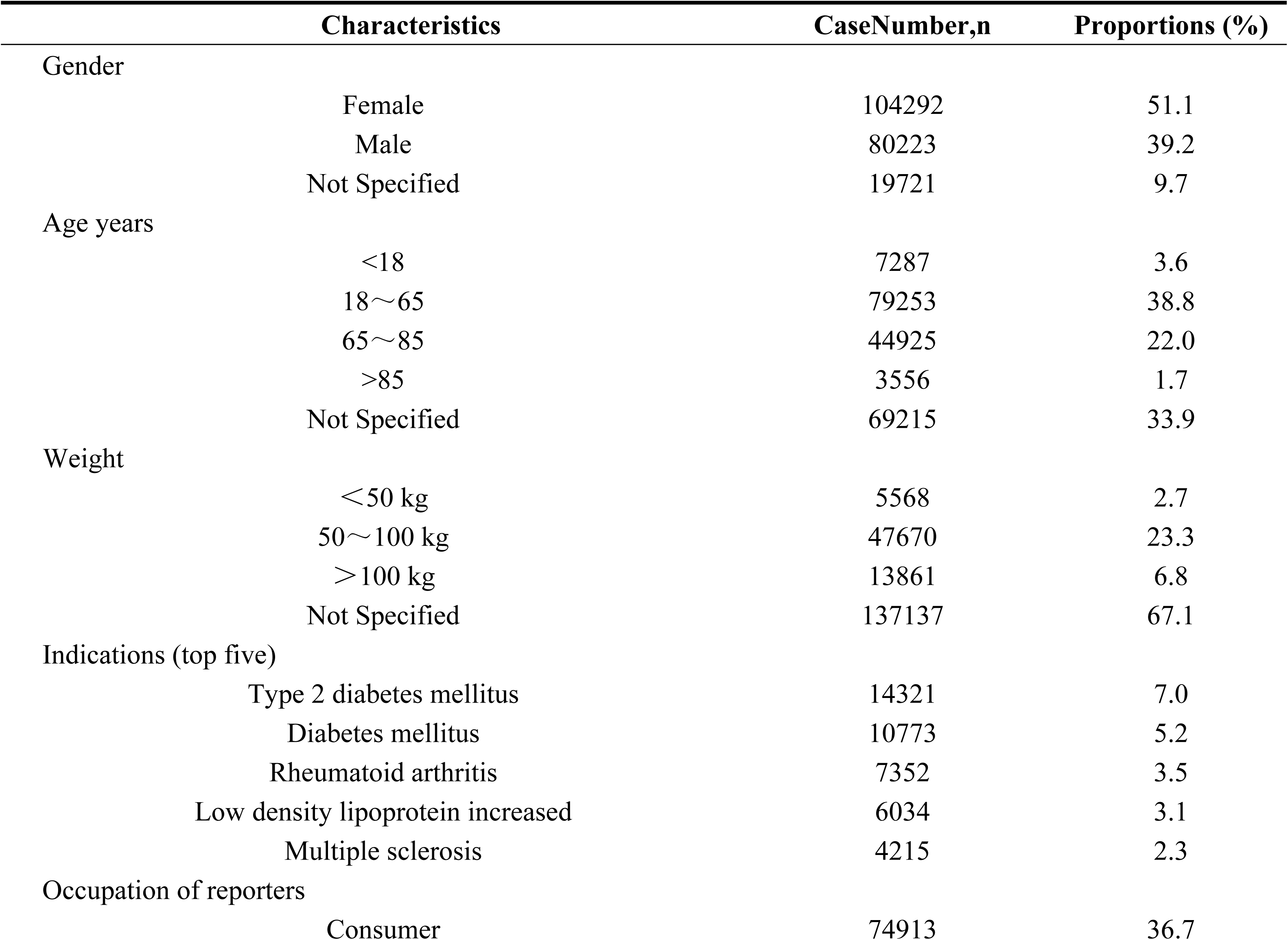
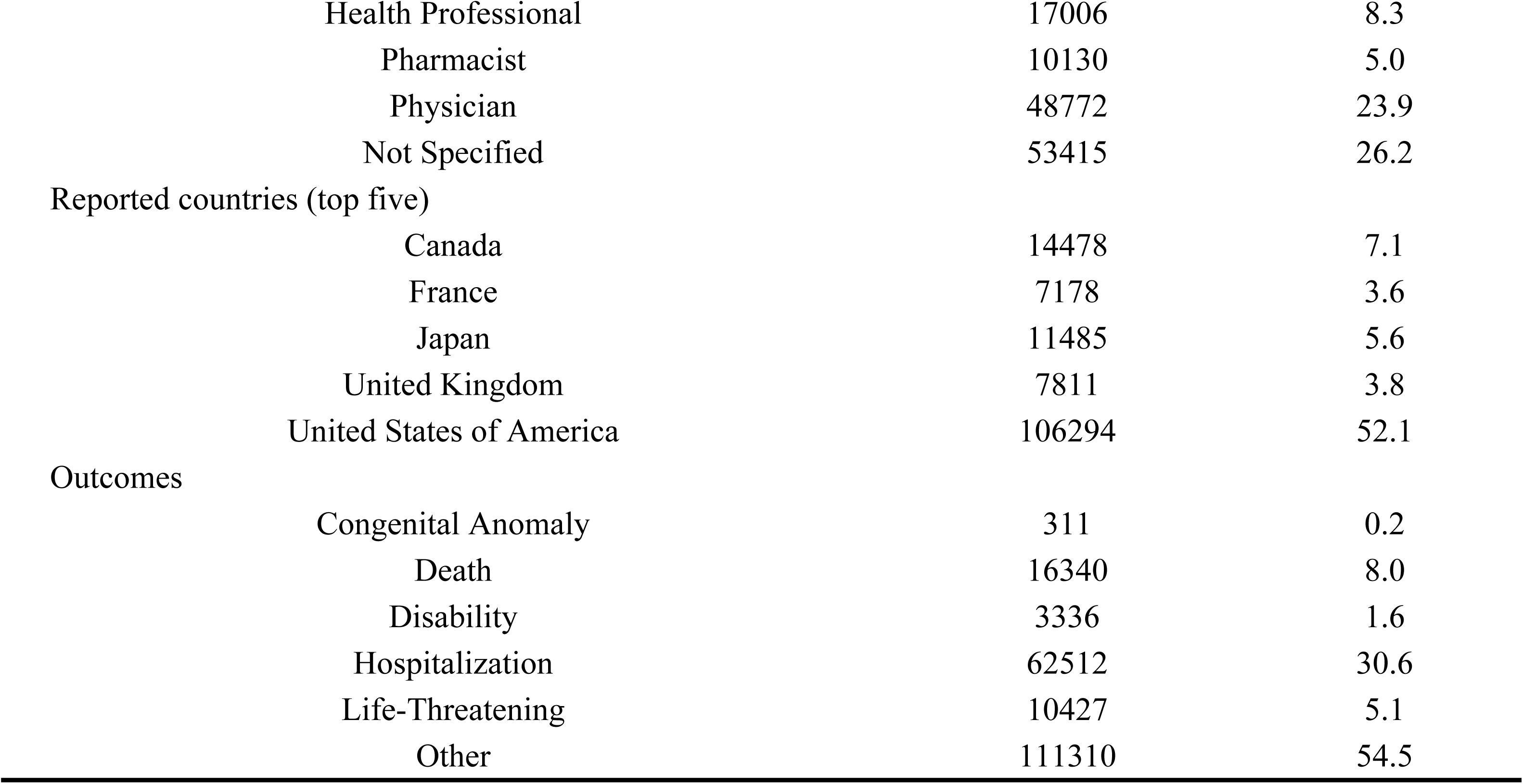
Analysis of basic patient data.

|  | Characteristics | CaseNumber,n | Proportions (%) |
| --- | --- | --- | --- |
| Gender | Female | 104292 | 51.1 |
|  | Male | 80223 | 39.2 |
|  | Not Specified | 19721 | 9.7 |
| Age years | <18 | 7287 | 3.6 |
|  | 18~65 | 79253 | 38.8 |
|  | 65~85 | 44925 | 22.0 |
|  | >85 | 3556 | 1.7 |
|  | Not Specified | 69215 | 33.9 |
| Weight | <50 kg | 5568 | 2.7 |
|  | 50~100 kg | 47670 | 23.3 |
|  | >100 kg | 13861 | 6.8 |
|  | Not Specified | 137137 | 67.1 |
| Indications (top five) | Type 2 diabetes mellitus | 14321 | 7.0 |
|  | Diabetes mellitus | 10773 | 5.2 |
|  | Rheumatoid arthritis | 7352 | 3.5 |
|  | Low density lipoprotein increased | 6034 | 3.1 |
|  | Multiple sclerosis | 4215 | 2.3 |
| Occupation of reporters | Consumer | 74913 | 36.7 |

|  |  |  |
| --- | --- | --- |
| Health Professional | 17006 | 8.3 |
| Pharmacist | 10130 | 5.0 |
| Physician | 48772 | 23.9 |
| Not Specified | 53415 | 26.2 |
| Reported countries (top five) |  |  |
| Canada | 14478 | 7.1 |
| France | 7178 | 3.6 |
| Japan | 11485 | 5.6 |
| United Kingdom | 7811 | 3.8 |
| United States of America | 106294 | 52.1 |
| Outcomes |  |  |
| Congenital Anomaly | 311 | 0.2 |
| Death | 16340 | 8.0 |
| Disability | 3336 | 1.6 |
| Hospitalization | 62512 | 30.6 |
| Life-Threatening | 10427 | 5.1 |
| Other | 111310 | 54.5 |

The yearly trend of adverse drug events reports related to glucose metabolism disorders is illustrated in Fig 3, revealing that the highest number of cases was reported in 2015. After searching for the HLT contained in “Glucose metabolism disorders,” A total of 45 related Preferred Terms were identified and were detailed in S3 Table. The top 30 PTs, ranked by drug proportion, were listed in Table 2.

**Fig 3.**
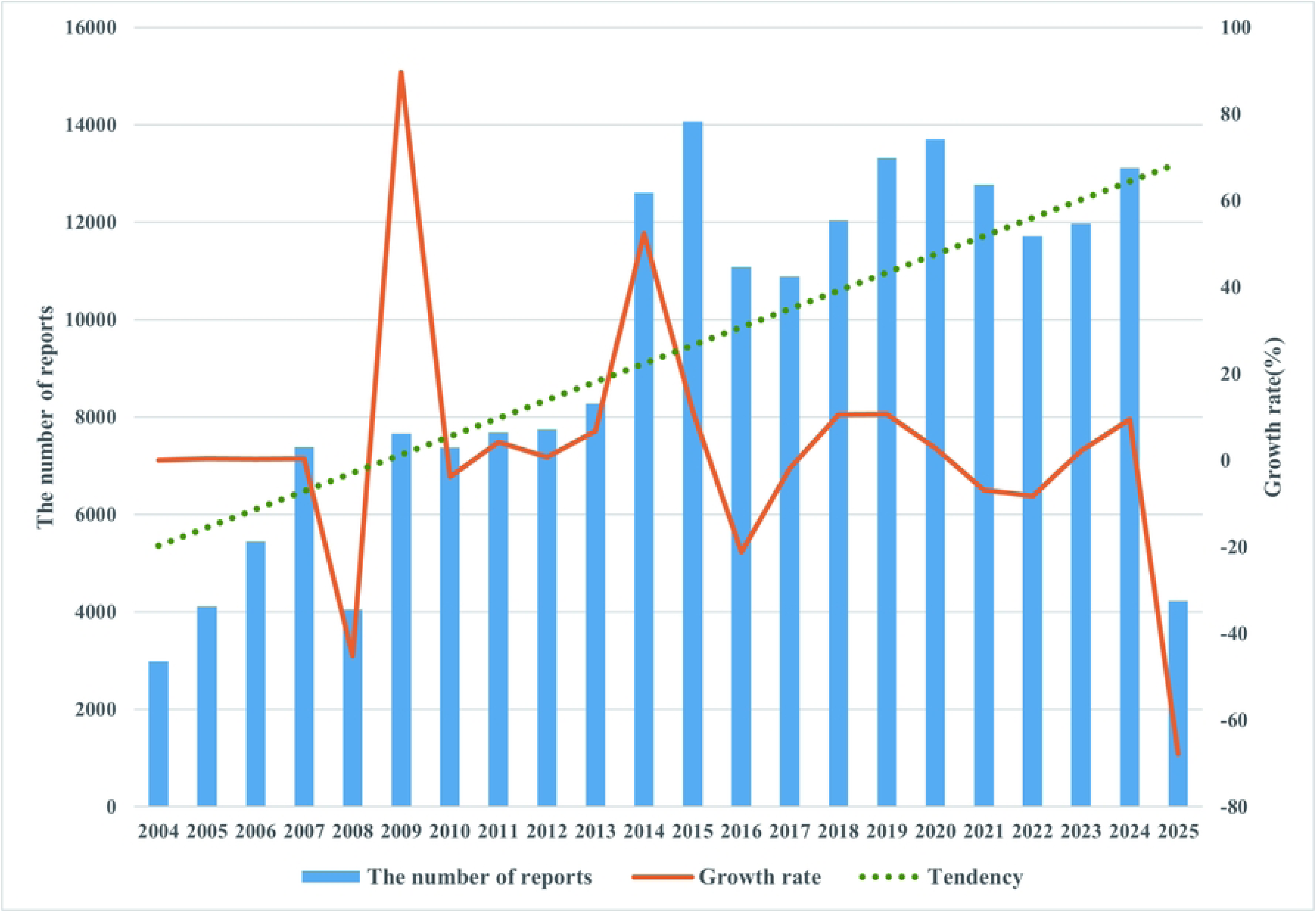
Annual trend in reporting of adverse drug events related tc.

**Table 2.**
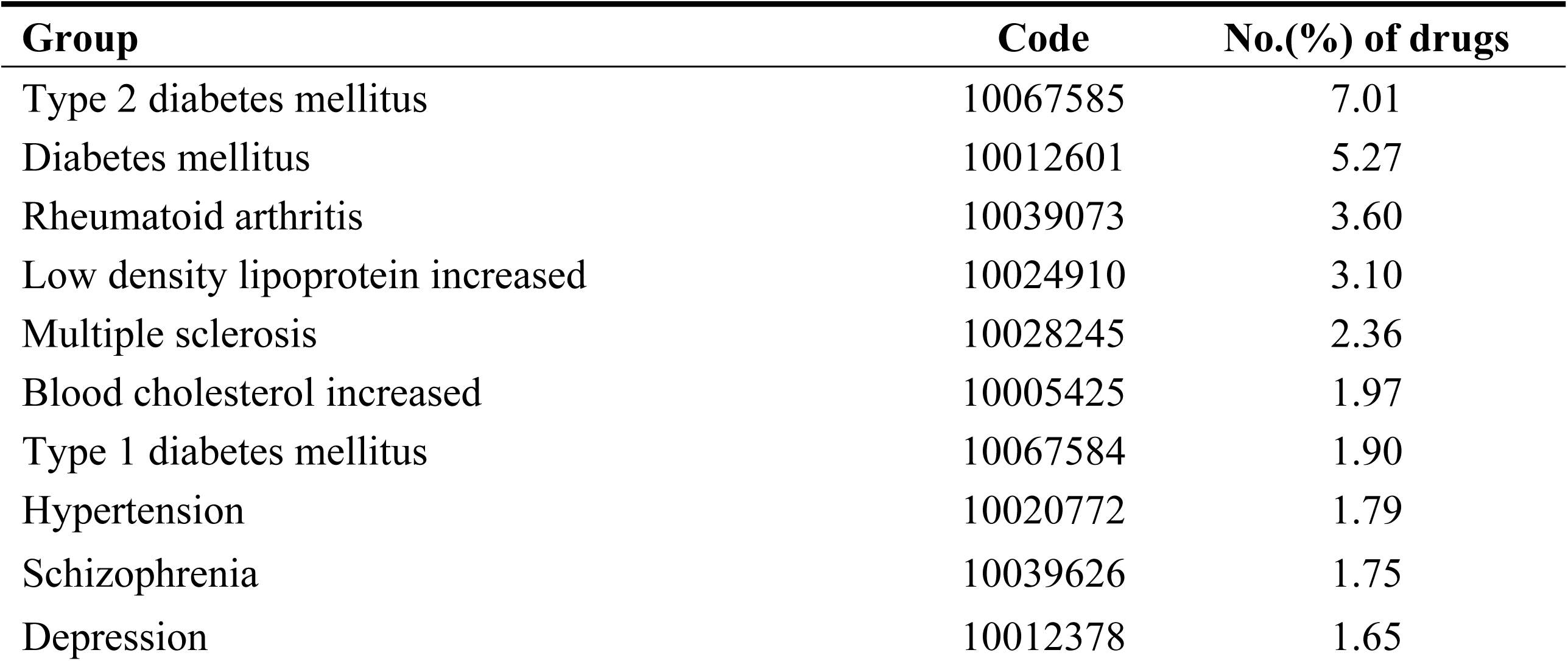
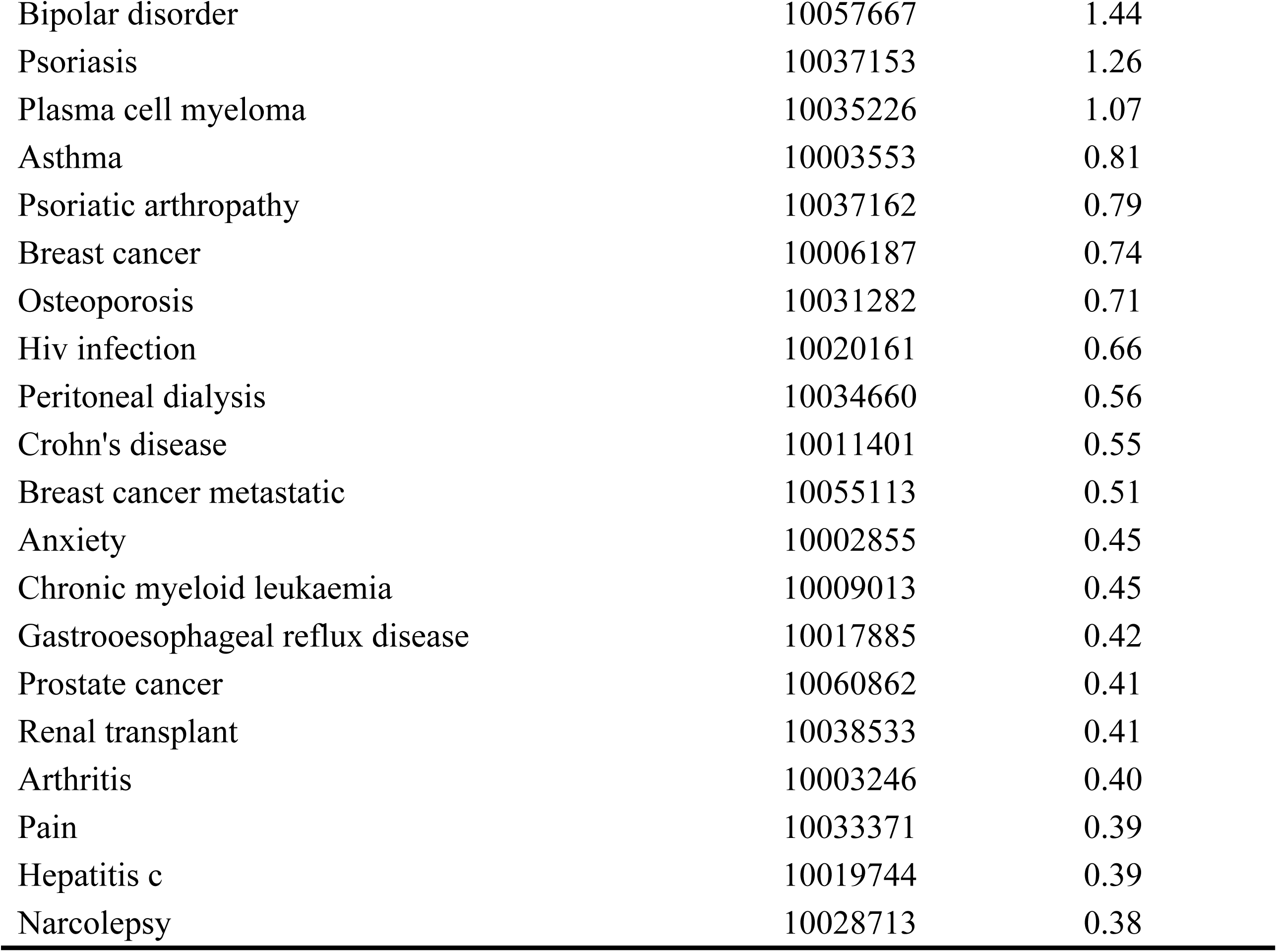
Number of drugs associated with PT group(top 30)

| Group | Code | No.(%) of drugs |
| --- | --- | --- |
| Type 2 diabetes mellitus | 10067585 | 7.01 |
| Diabetes mellitus | 10012601 | 5.27 |
| Rheumatoid arthritis | 10039073 | 3.60 |
| Low density lipoprotein increased | 10024910 | 3.10 |
| Multiple sclerosis | 10028245 | 2.36 |
| Blood cholesterol increased | 10005425 | 1.97 |
| Type 1 diabetes mellitus | 10067584 | 1.90 |
| Hypertension | 10020772 | 1.79 |
| Schizophrenia | 10039626 | 1.75 |
| Depression | 10012378 | 1.65 |
| Bipolar disorder | 10057667 | 1.44 |
| Psoriasis | 10037153 | 1.26 |
| Plasma cell myeloma | 10035226 | 1.07 |
| Asthma | 10003553 | 0.81 |
| Psoriatic arthropathy | 10037162 | 0.79 |
| Breast cancer | 10006187 | 0.74 |
| Osteoporosis | 10031282 | 0.71 |
| Hiv infection | 10020161 | 0.66 |
| Peritoneal dialysis | 10034660 | 0.56 |
| Crohn's disease | 10011401 | 0.55 |
| Breast cancer metastatic | 10055113 | 0.51 |
| Anxiety | 10002855 | 0.45 |
| Chronic myeloid leukaemia | 10009013 | 0.45 |
| Gastrooesophageal reflux disease | 10017885 | 0.42 |
| Prostate cancer | 10060862 | 0.41 |
| Renal transplant | 10038533 | 0.41 |
| Arthritis | 10003246 | 0.40 |
| Pain | 10033371 | 0.39 |
| Hepatitis c | 10019744 | 0.39 |
| Narcolepsy | 10028713 | 0.38 |

Fig 4 illustrates the duration from the initiation of medication to the onset of related adverse events. The median time to onset was 108 days (IQR 8 – 553), with 50% of cases occurring within the first 100 days. Additionally, approximately 25% of reports were noted to occur between 100 and 550 days.

**Fig 4.**
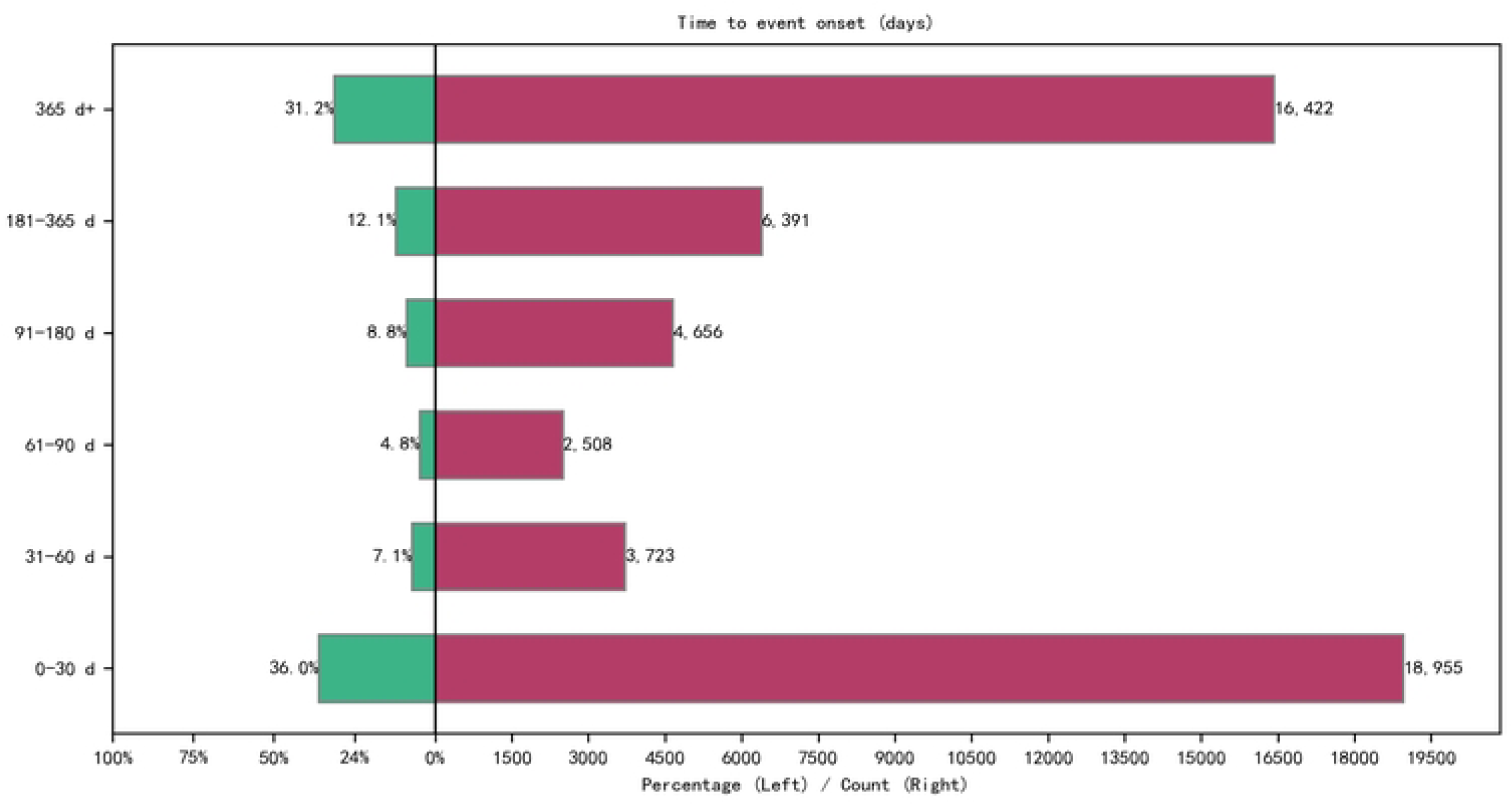
The time interval between drug use and the onset of drug.

### 3.2 Drugs associated with an increased the risk of glucose metabolism disorders

To facilitate international comparison and interpretation by drug class, we assessed the risk signals associated with the top 30 drugs causing glucose metabolism disorders by categorizing them according to the Anatomical Therapeutic Chemical (ATC) classification system (second level). The analysis included ten categories: drugs for diabetes treatment, systemic corticosteroids, antineoplastic agents, psycholeptics, lipid-modifying agents, agents acting on the renin-angiotensin system, systemic antibacterials, anti-inflammatory and antirheumatic products, drugs for bone disease treatment, and pituitary and hypothalamic hormones and analogues. As shown in Fig 5, drugs related to type 2 diabetes constitute 7.01% and were classified under the diabetes treatment category. Drugs associated with rheumatoid arthritis represented 3.60% and were classified as anti-inflammatory and antirheumatic products. Additionally, drugs linked to hypertension accounted for 1.79% and were categorized under agents acting on the renin-angiotensin system.

**Fig 5.**
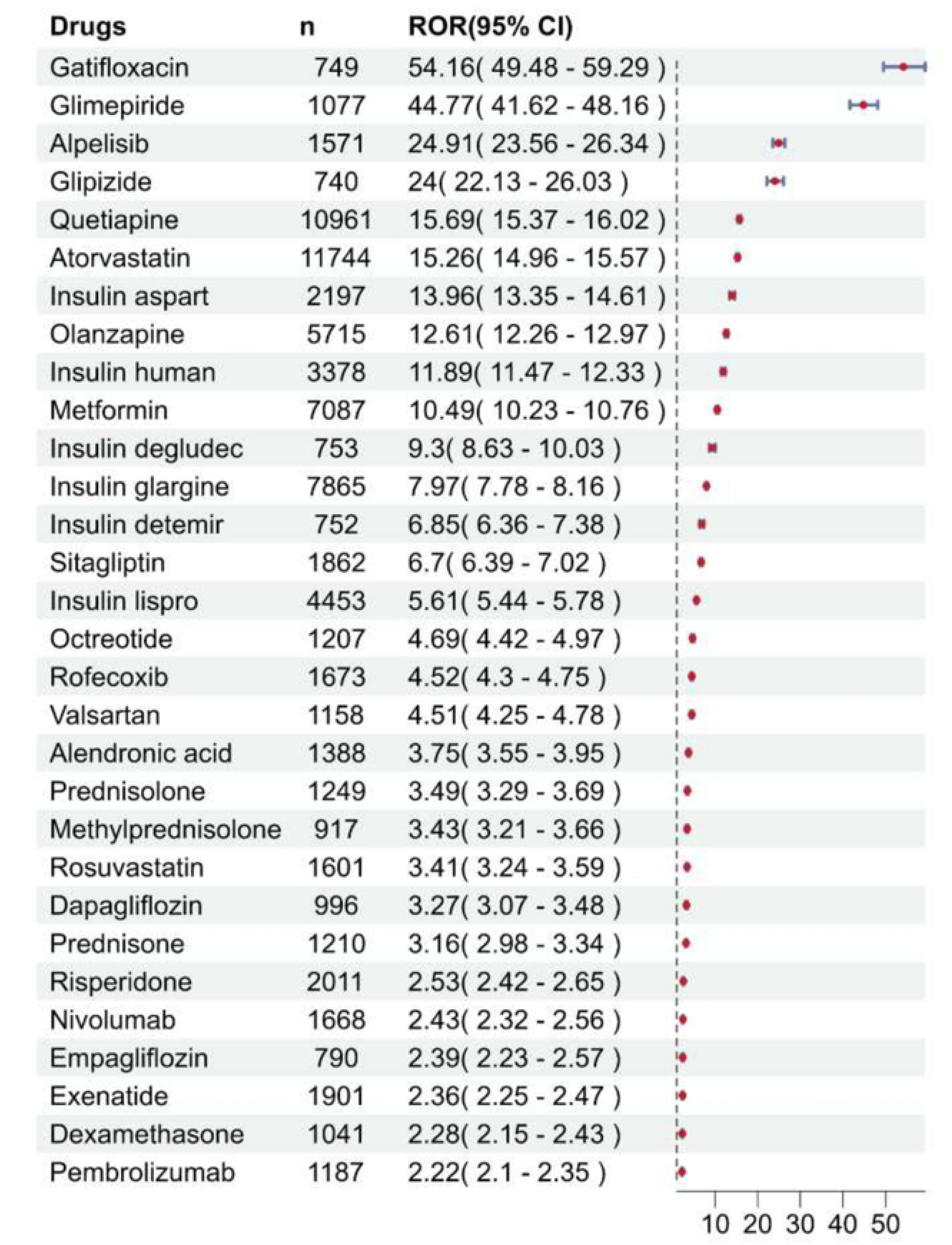
ROR for glucose metabolism disorders of s.

### 3.3 Single-drug risk signal detection

To further elaborate on the risk intensity associated with individual drugs, we could conclude that a total of 128 drugs exhibited positive signals. The point estimates and 95% confidence intervals for all four disproportionality metrics (ROR, PRR, EBGM, IC) for these 128 drugs are provided in S3 Table. The top 10 drugs with positive signals, ranked by their ROR signal strength, were as follows: Tofogliflozin [ROR (95% CI): 183.45 (33.6 – 1001.63)], amino acid compound preparation [ROR (95% CI): 63.06 (29.27 – 135.89)], repaglinide [ROR (95% CI): 60.6 (54.1 – 67.89)], gliclazide [ROR (95% CI): 58.71 (37.67 – 91.51)], gatifloxacin [ROR (95% CI): 54.16 (49.48 – 59.29)], glibenclamide [ROR (95% CI): 53.31 (48.46 – 58.65)], inavolisib [ROR (95% CI): 52.42 (25.79 – 106.54)], insulin nos [ROR (95% CI): 47.96 (39.07 – 58.89)], and acetylsalicylic acid aluminum glycinate magnesium carbonate [ROR (95% CI): 45.86 (13.81 – 152.31)]. A higher ROR value indicates a stronger risk signal, suggesting an increased risk of drug-associated glucose metabolism disorders. Consequently, a greater ROR value correlates with a higher likelihood of adverse events associated with the use of specific drugs(16).

## 4. Discussion

This disproportionality analysis identified 128 drugs with positive signals associated with glucose metabolism disorders in the FAERS database. This study strictly followed the FDA-recommended data cleaning process, which involved the removal of duplicate and invalid reports. The accuracy of drug classification was ensured by utilizing standardized drug names. Four algorithms were integrated to identify positive signals, significantly mitigating the risk of false positives and enhancing the reliability of the results. Furthermore, the study concentrated exclusively on ‘primary suspect drugs,’ thereby excluding potential interference from concomitant medications or underlying conditions, which further reinforced the directionality of causal inference.

As anticipated, anti-diabetic medications constituted the largest proportion of drugs with positive signals (38%), which is expected given their mechanism of action and the underlying condition of the patient population. However, the key findings of this analysis extend beyond known associations. We identified significant reporting associations for a diverse range of non-antidiabetic drugs, including antineoplastic agents (9.3%), agents acting on the renin-angiotensin system (8.6%), systemic corticosteroids (4.7%), and psycholeptics (3.9%). This highlights the broader potential for pharmacologically diverse agents to disrupt glucose homeostasis. Notably, while signals for anti-diabetic drugs and systemic corticosteroids were expected based on known mechanisms, signals for certain antineoplastic agents (e.g., enfortumab vedotin) and other drug classes represent potentially novel associations or highlight underappreciated risks not adequately reflected in current drug labels.

Among the available anthropometric data on body weight, the majority of reported cases were distributed in the 50 – 100 kg range (23.3%), followed by the >100 kg subgroup (6.8%). However, a critical data gap was observed, with body weight information missing in 67.1% of reports, representing a substantial limitation for elucidating the interplay between body weight and drug-induced glucose metabolism disorders. Obesity has long been recognized as a pivotal risk factor for glucose metabolism disorders(18). Accumulating evidence indicates that obesity-induced insulin resistance, rooted in chronic low-grade adipose tissue inflammation and adipocyte dysfunction, serves as a central pathogenic mechanism that disrupts glucose homeostasis(19). Furthermore, obesity-driven ectopic lipid deposition in the hepatic and skeletal muscle tissues aggravates insulin resistance, thereby exacerbating disturbances in glucose metabolism(20). The limited availability of weight data in this study suggests that the regulatory role of obesity, a well-established risk factor for insulin resistance, may be underestimated in real-world clinical contexts. Given the profound impact of obesity on glucose metabolism, clinicians should emphasize the systematic collection and longitudinal monitoring of patients’ weight metrics, particularly when prescribing pharmacotherapies with potential metabolic side effects. This approach facilitates more precise risk stratification of drug-induced glucose metabolism disorders and the formulation of individualized therapeutic strategies.

The United States (n = 106,294; 52.1%) and Canada (n = 14,478; 7.1%) reported the highest number of cases, followed by Japan (n = 11,485; 5.6%). The higher reporting rates in the U.S. and Canada may be attributed to the prevalence of conditions predisposing individuals to glucose metabolism disorders. In the U.S., 11.6% of the population was diagnosed with diabetes in 2021(21). Additionally, high-sugar and high-fat dietary patterns contribute to the development of diabetes and related metabolic disorders(22, 23), which in turn increases the population’s susceptibility to drug-induced glucose disturbances, leading to a rise in related reports. In Canada, approximately 9% of adults aged 20 to 79 were diagnosed with diabetes between 2016 and 2019(^24^), with rates reaching 18% among those aged 60 to 79(^25^). Dietary and lifestyle factors contributing to this prevalence may similarly elevate the risk of drug-induced glucose metabolism disorders, thereby increasing the number of reports(26). Despite dietary patterns differing from the US and Canada, an aging population is likely to contribute to higher diabetes prevalence in Japan(27). Research indicated that in 2019, the overall prevalence rate of diabetes in Japan was 5.6%, with the risk of developing the disease escalating with age(28). This elevated prevalence of diabetes and associated glucose metabolism disorders, exacerbated by Japan’s aging demographic, may explain the relatively high number of reports from the country, as a greater number of patients with underlying metabolic issues are exposed to medications that disrupt glucose homeostasis(29).

Among the 128 positive signal drugs, those used for diabetes treatment constituted a significantly higher proportion compared to other drug categories, accounting for 38%. This finding suggested a strong association between these medications and glucose metabolism disorders. Other drug categories included antineoplastic agents (9.3%), agents acting on the renin-angiotensin system (8.6%), corticosteroids for systemic use (4.7%), and pituitary and hypothalamic hormones and analogues (4.7%), as well as psycholeptics (3.9%). Furthermore, the study revealed that certain compound formulations, such as antacids containing aluminum and magnesium, also displayed higher signals. This indicated that non-single components formulations may disrupt glucose homeostasis through synergistic effects.

Similarly, the proportion of diabetes medications utilized is significantly higher than that of other pharmaceuticals, primarily due to the following reasons: First, the population utilizing diabetes medications inherently exhibits deficiencies in pancreatic beta cell function and insulin resistance, resulting in a markedly elevated pre-existing risk of hyperglycemia or hypoglycemia compared to the general population(30–32). Even with adherence to prescribed regimens, fluctuations in blood glucose levels may be attributed to ‘adverse drug reactions’ leading to an inflated number of reports in the FAERS. Results from an international multicenter observational study involving over 27,000 diabetes patients treated with insulin (including both Type 1 and Type 2 Diabetes Mellitus) indicated that, under standardized dose adjustment protocols, 24.8% of Type 2 Diabetes Mellitus patients experienced at least one episode of symptomatic hypoglycemia within the past four weeks; only approximately one-third (34%) of these cases were clinically confirmed to be related to insulin dosage, while the remainder were attributed to factors such as irregular eating patterns, excessive physical activity, or variations in insulin sensitivity(33). Secondly, there is an ‘amplification effect’ in reporting behavior. Diabetic patients are required to routinely monitor their blood glucose levels, making them more likely to detect asymptomatic hyperglycemia or hypoglycemia and subsequently report these events. In contrast, non-diabetic patients using glucocorticoids may overlook abnormal blood glucose levels unless symptoms manifest or hospitalization occurs(34).

A substantial body of real-world data, particularly reports from drug vigilance databases such as FAERS and EudraVigilance, identifies anticancer drugs as the second largest class of medications responsible for adverse effects related to hyperglycemia or hypoglycemia, following insulin and insulin secretagogues(7, 35, 36). Numerous mechanistic studies have investigated the relationship between antineoplastic agents and glucose metabolism-related adverse events. For instance, targeted kinase inhibitors, such as phosphatidylinositol 3-kinase (PI3K)/Protein Kinase B (AKT)/mammalian target of rapamycin (mTOR) inhibitors like alpelisib and capivasertib, can disrupt insulin signaling pathways and inhibit pancreatic β-cell proliferation, resulting in insulin-deficient hyperglycemia(37–39). BCR-ABL tyrosine kinase inhibitors suppress the phosphorylation of insulin receptor substrate-1 and promote the expression of gluconeogenesis-related genes, leading to steroid-like diabetes(40–43). Immune checkpoint inhibitors (ICIs), such as nivolumab and pembrolizumab may cause significant disturbances in glucose homeostasis due to their off-target immune activation. A comprehensive review of relevant studies has proposed a dual-pathway model in which ICIs simultaneously activate anti-β-cell and anti-pituitary immune responses, resulting in a range of glucose disorders from acute insulin deficiency-induced hyperglycemia to Adrenocorticotropic Hormone (ACTH) deficiency-induced hypoglycemia(44–49). Early identification and timely hormone replacement therapy are crucial for mitigating life-threatening metabolic crises.

It is worth noting that our analysis has revealed a critical oversight: many anti-cancer drugs that may cause drug-induced glucose metabolism disorders, such as enfortumab vedotin and mercaptopurine, do not have warnings about such metabolic risks in their drug instructions. This information gap constitutes a significant deficiency for healthcare providers and patients alike. Given that drug-induced glucose metabolism disorders can exacerbate the conditions of both diabetic and cancer patients and may complicate cancer treatment, our findings highlight the urgent need for more comprehensive safety information on anti-cancer drugs. Moreover, as new anti-cancer drugs continue to emerge, especially those innovative therapies that are transforming the landscape of cancer treatment, it is crucial to continuously monitor the issue of glucose metabolism disorders caused by such drugs.

We observed that among the positive signals associated with agents acting on the renin-angiotensin system, hydrochlorothiazide and its combination drugs accounted for the majority, representing 36%. Our findings were consistent with the widely accepted view that thiazide diuretics may impair glucose metabolism(50–52). Research indicated that thiazide diuretics, such as hydrochlorothiazide, exerted a direct toxic effect on pancreatic beta cells(53–55). Prolonged exposure to these agents compromised the normal functioning of beta cells, resulting in reduced insulin synthesis and secretion. Additionally, thiazide diuretics promote potassium excretion, frequently leading to hypokalemia(56). Notably, the normal function of pancreatic beta cells relies on the maintenance of adequate intracellular potassium levels, which are essential for the regulation of ion channels and membrane potential Hypokalemia disrupts this critical balance, thereby contributing to hyperglycemia(57). Another drug targeting the renin-angiotensin system, captopril, was found in this study to be associated with signals related to glucose metabolism disorders, and previous studies have also reported similar risks(58, 59). However, the risk is not explicitly mentioned in the drug’s instructions.

In this analysis, systemic corticosteroids, pituitary and hypothalamic hormones and their analogues, exhibited positive associations with glucose metabolism disorders, which was consistent with findings from previous studies. Systemic corticosteroids induce metabolic disorders through various mechanisms, including: (1) promoting hepatic gluconeogenesis, thereby increasing glucose production; (2) inhibiting peripheral glucose uptake and utilization, resulting in elevated blood glucose levels; (3) enhancing insulin resistance, thereby exacerbating hyperglycemia due to reduced insulin sensitivity; and (4) interfering with pancreatic β-cell function, thereby affecting normal insulin secretion(10, 60, 61). Pasireotide, a relatively new somatostatin analogue, demonstrates a higher affinity for SSTR5, approximately 40 times greater than that of other analogues. By targeting SSTR5, it significantly inhibits insulin secretion, leading to elevated blood glucose levels. Furthermore, pasireotide may also influence the secretion of glucagon-like peptide-1 (GLP-1), thereby increasing the risk of hyperglycemia(62, 63). In contrast, octreotide and lanreotide primarily exert their effects by binding to SSTR2 on pancreatic β-cells, which inhibits adenylyl cyclase activity. This inhibition subsequently reduces intracellular cyclic adenosine monophosphate (cAMP) levels and blocks voltage-gated calcium channels, ultimately suppressing insulin secretion(64, 65).

With the class of psycholeptics, certain drugs are associated with an increased risk of glucose metabolism disorders. These include atypical antipsychotics such as quetiapine, olanzapine, and risperidone, as well as the typical antipsychotic fluphenazine Among them, olanzapine and quetiapine may cause weight gain and visceral fat accumulation by blocking multiple receptors, which in turn causes insulin resistance and directly impairs pancreatic β-cell function. This functional impairment inhibits insulin secretion and reduces glucose sensitivity, thereby significantly increasing the risk of metabolic disorders(66–69). In contrast, risperidone and fluphenazine demonstrate comparatively milder metabolic effects. Risperidone may influence glucose metabolism indirectly through long-term hyperprolactinemia, while fluphenazine exerts mild anticholinergic effects that weakly reduce insulin secretion(70–73).

From a clinical perspective, these findings suggest that healthcare providers should consider enhanced glucose monitoring in patients receiving high-risk medications, particularly those with existing metabolic risk factors or those using drugs with newly identified signals (e.g., certain antineoplastic agents). Timely recognition and management of drug-induced dysglycemia may improve patient safety and treatment outcomes.

This study utilized data from the FAERS database, a large-scale spontaneous reporting system for adverse drug reactions maintained by the U.S. FDA. It offers significant advantages, including extensive coverage, a substantial data volume, and timely updates. This database can capture potential safety issues related to various drugs in clinical applications worldwide in real time, thereby providing ample real-world data support for pharmacovigilance research. However, the utility of FAERS in pharmacovigilance is constrained by several inherent methodological limitations. Most importantly, it is crucial to emphasize that disproportionality analyses, as performed in this study, detect reporting associations that do not, in themselves, prove causation. First, the spontaneous reporting design does not facilitate robust causal inference due to the absence of reliable exposure denominators and the inability to control for confounding factors such as polypharmacy and comorbidities. (No sensitivity analyses to control for confounding, e.g., through stratification, were performed in this study, which is a limitation). Second, data heterogeneity, which includes incomplete or missing data fields (e.g., weight in 67.1% of reports) and differential reporting biases, undermines the stability and reliability of detected signals. Third, the lack of denominator data prevents accurate estimation of adverse event incidence rates, necessitating reliance on disproportionality metrics like the Reporting Odds Ratio, which are vulnerable to both false positives and false negatives. Fourth, FAERS data mainly comes from high-income countries, which limits the universality of its research conclusions and makes it difficult to apply them to populations of different economic levels or ethnicities around the world. Fifth, the absence of structured data on dose-response relationships and temporal sequences hinders the understanding of underlying biological mechanisms. The signals detected in this study represent statistical associations that require validation through robust epidemiological studies to assess causality and quantify risk. Despite these limitations, FAERS remains a valuable large-scale real-world repository for hypothesis generation in pharmacovigilance.Furthermore, the spontaneous reporting design and lack of exposure denominators mean that disproportionality analyses cannot prove causation or measure the incidence of adverse events.

## 5. Conclusion

This study systematically analyzed data from the FAERS database (Q4 2004–Q1 2025) employing four disproportionality analysis methods, identifying 128 drugs associated with positive signals for glucose metabolism disorders. These drugs primarily include anti-diabetic medications (38%), antineoplastic agents (9.3%), systemic corticosteroids, agents acting on the renin-angiotensin system, and psycholeptics. Mechanistically, they disrupt glucose homeostasis through various pathways such as the induction of insulin resistance, impairment of pancreatic β-cell function, and interference with immune regulatory processes.

Notably, there are significant gaps in drug labeling. The antineoplastic agents (such as enfortumab vedotin and mercaptopurine) that have been confirmed positive signals, lack explicit warnings regarding glucose metabolism disorders. Given the potential risks of exacerbating metabolic conditions in vulnerable populations (e.g., cancer or diabetes patients), it is crucial to update drug labels and strengthen monitoring of high-risk drugs, particularly newly emerging anticancer agents. In addition, one limitation of this study is the absence of weight data in 67.1% of reports, which may lead to an underestimation of the role of obesity.This highlights the necessity for future research that incorporates comprehensive patient-level information. The signals detected in this hypothesis-generating study require further validation through robust epidemiological studies to assess causality and quantify risk.

In conclusion, this disproportionality analysis identified reporting associations for 128 drugs with glucose metabolism disorders, primarily anti-diabetic, antineoplastic, and cardiovascular agents. It is crucial to emphasize that these findings are hypothesis-generating. The significant gaps in drug labeling for certain antineoplastic agents highlight an area requiring further investigation and regulatory attention. The limitations of this study, particularly the missing weight data and inability to control for confounding, underscore the necessity for future research that incorporates comprehensive patient-level information to validate these signals and assess their clinical relevance.

## Data availability statement

The data underlying this article were derived from publicly available sources: the FAERS database (https://fis.fda.gov/extensions/FPD-QDE-FAERS/FPD-QDE-FAERS.html). The derived data generated in this research (the list of positive signals with disproportionality metrics) will be shared on reasonable request to the corresponding author.

## Code availability statement

The custom SAS code used for data cleaning and disproportionality analysis is available from the corresponding author upon reasonable request. Software used: SAS 9.4, Navicat 16, Microsoft Excel 2021, SPSS 27.0.1, GraphPad Prism 9.5.

## Ethics approval

Ethics approval was not required for this study as it utilized only publicly available, anonymized data.

## Informed consent

Informed consent was not required for this study due to its retrospective design and use of anonymized data.

## Abbreviations

ACTH: Adrenocorticotropic Hormone
AEs: Adverse Events
AKT: Protein Kinase B
ATC: Anatomical Therapeutic Chemical
BCPNN: Bayesian Confidence Propagation Neural Network
cAMP: Cyclic Adenosine Monophosphate
EBGM: Empirical Bayes Geometric Mean
FAERS: FDA Adverse Event Reporting System
GLP-1: Glucagon-like Peptide-1
HLT: High-Level Term
IC025: Information Component 025
ICIs: Immune Checkpoint Inhibitors
MGPS: Multi-item Gamma Poisson Shrinker
MedDRA: Medical Dictionary for Regulatory Activities
mTOR: Mammalian Target of Rapamycin
PI3K: Phosphatidylinositol 3-kinase
PRR: Proportional Reporting Ratio
PS: Primary Suspected Drug
PT: Preferred Term
ROR: Reporting Odds Ratio
SSTR5: Somatostatin Receptor 5
T1DM: Type 1 Diabetes Mellitus
T2DM: Type 2 Diabetes Mellitus

## Acknowledgements

Thanks to the US FDA for providing the data source of this study.

## Author contributions

Zhongxiang Zhang: Data curation, Methodology, Resources, Software, Visualization, Writing–original draft, Writing–review and editing. Qiaoying Li: Data curation, Methodology, Writing–original draft, Writing–review and editing. Tao Huang: Conceptualization, Methodology, Supervision, Visualization, Writing–original draft. Xuping Yang: Conceptualization, Methodology, Software, Supervision. Xunyun Du: Conceptualization, Methodology, Software, Supervision. Xinyi Deng: Software,Visualization, Writing–original draft. Shurong Wang: Data curation, Methodology, Resources, Visualization, Writing–review and editing. Jie Zhou: Data curation, Methodology, Resources, Software, Visualization, Writing–original draft, Writing–review and editing.

---

**S1 Table.**
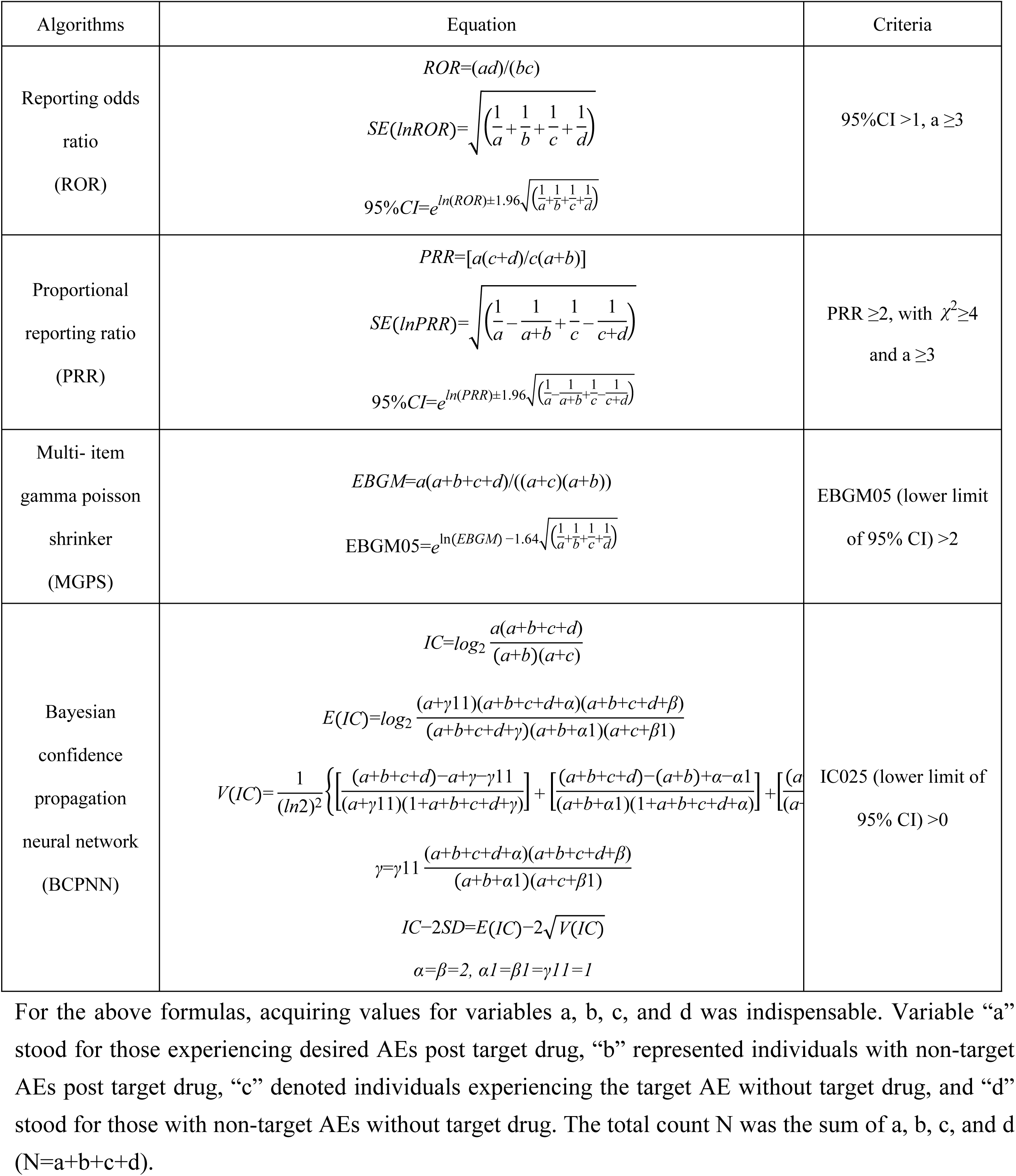
Four methods of disproportionality analysis.

**S2 Table.**
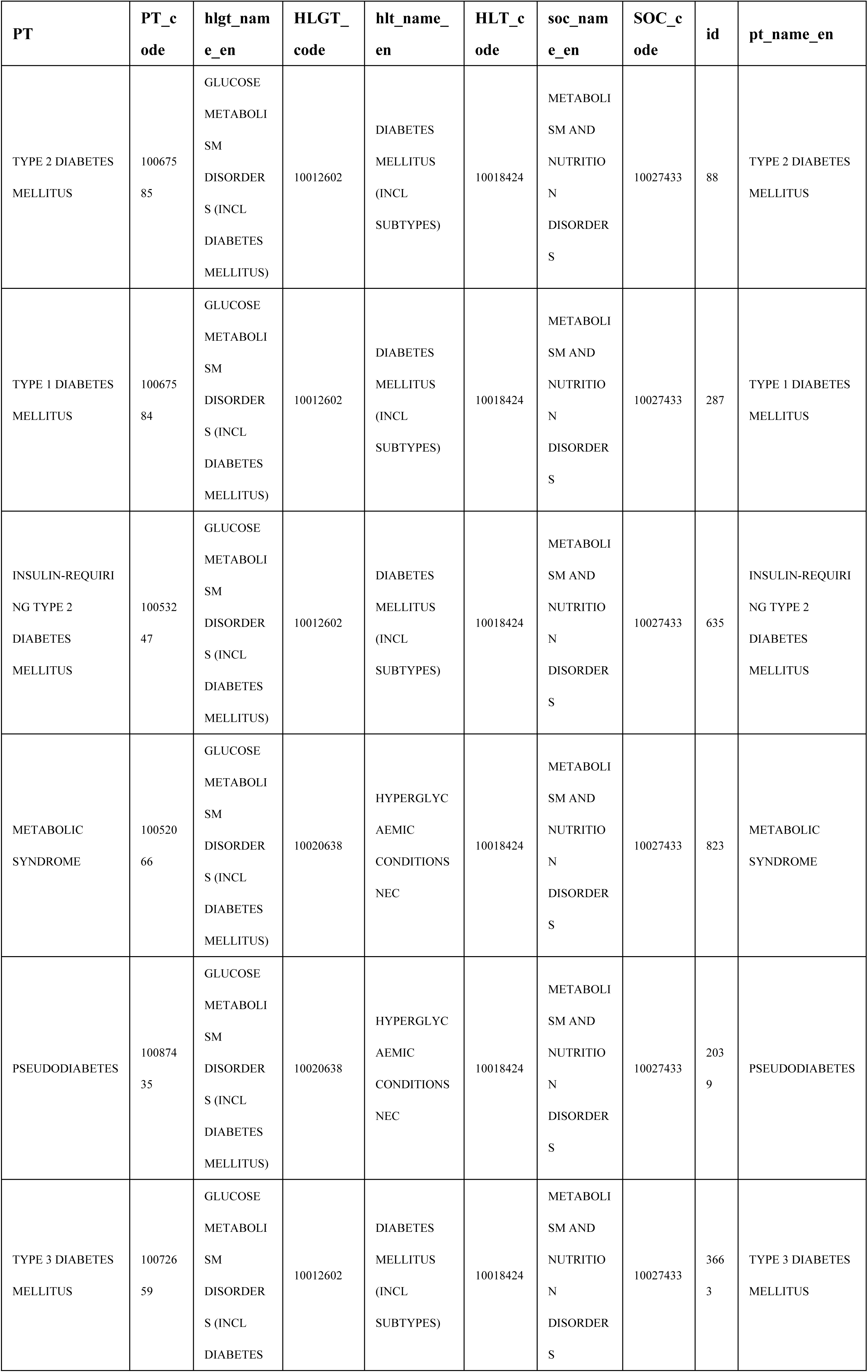
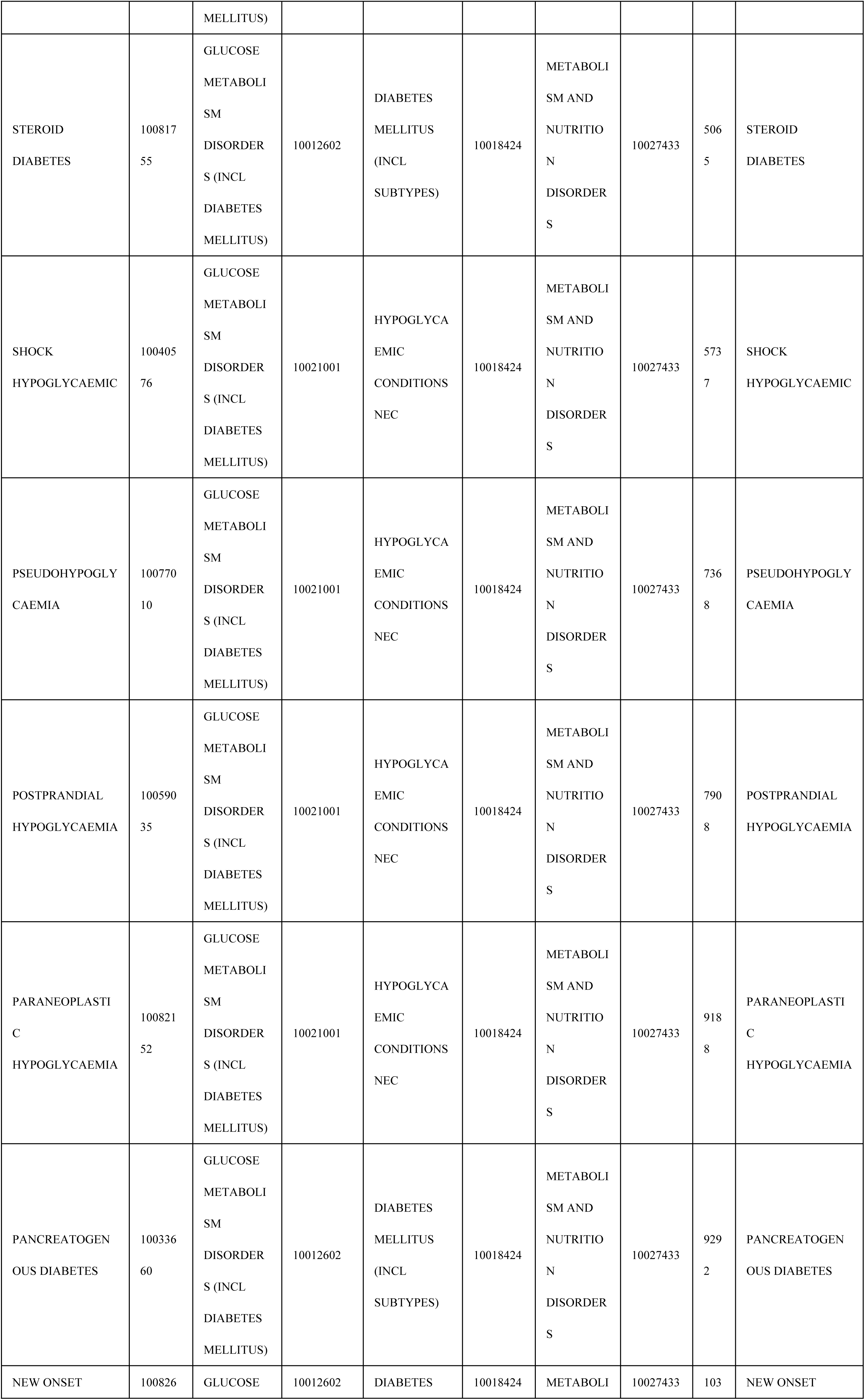
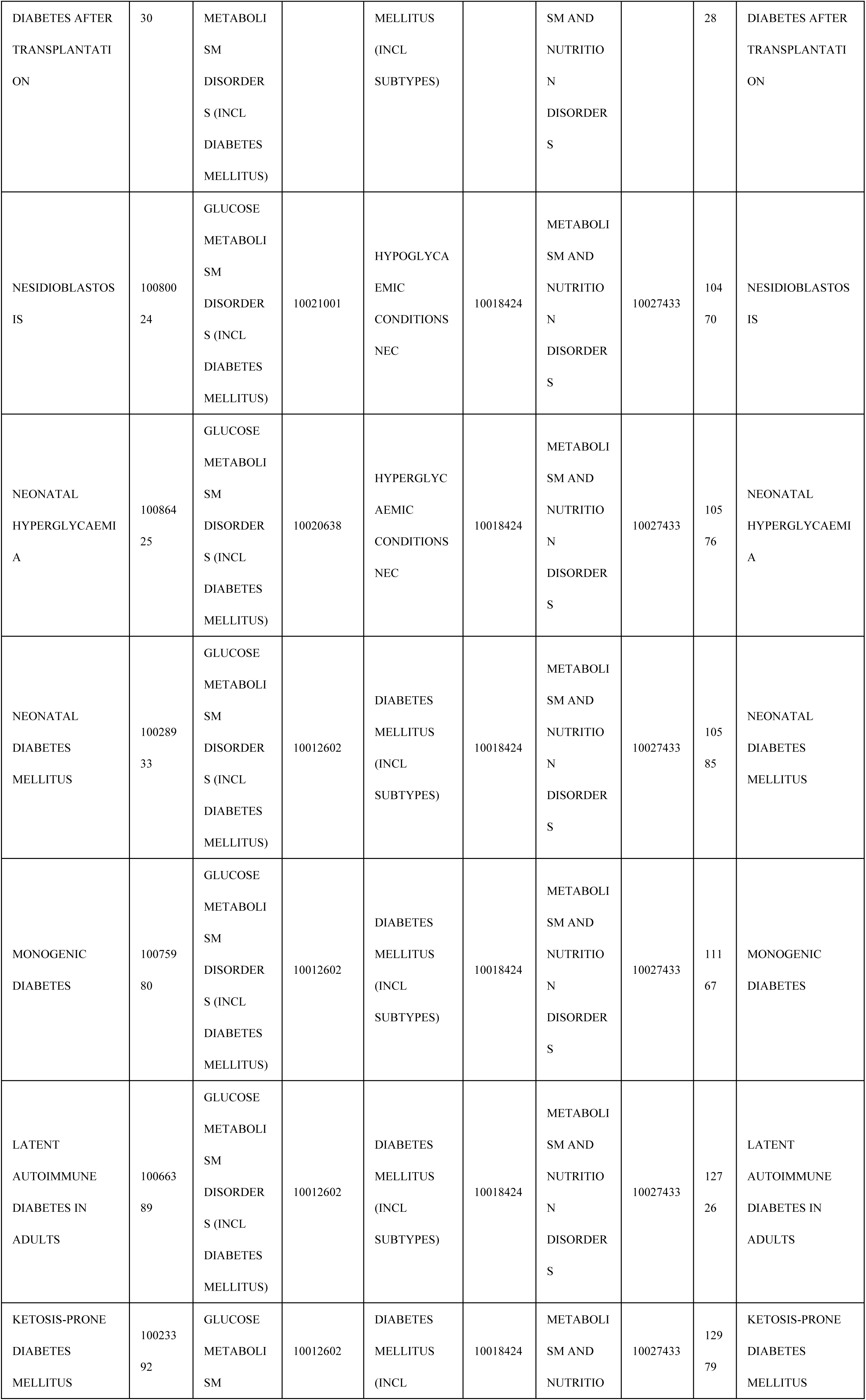
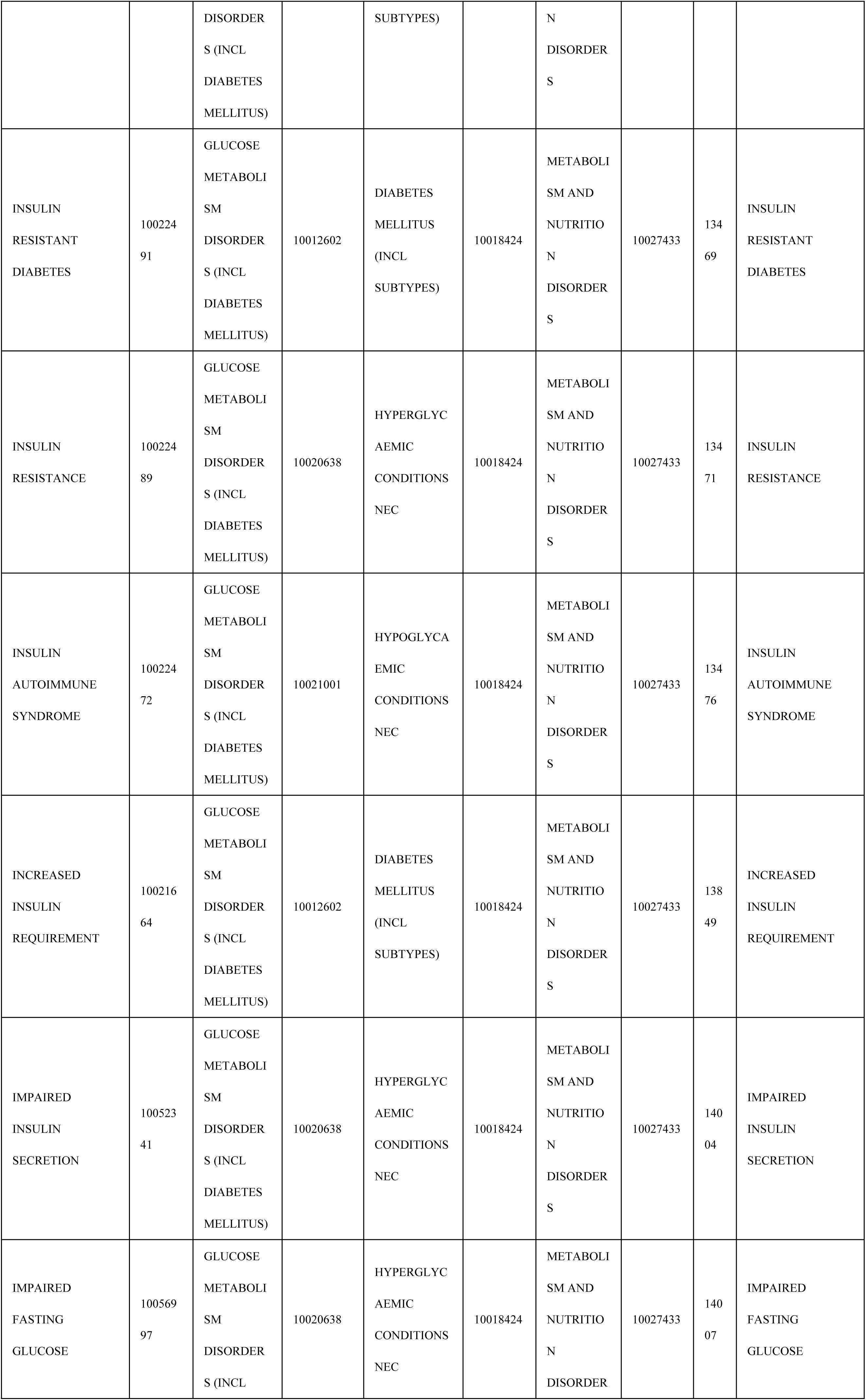
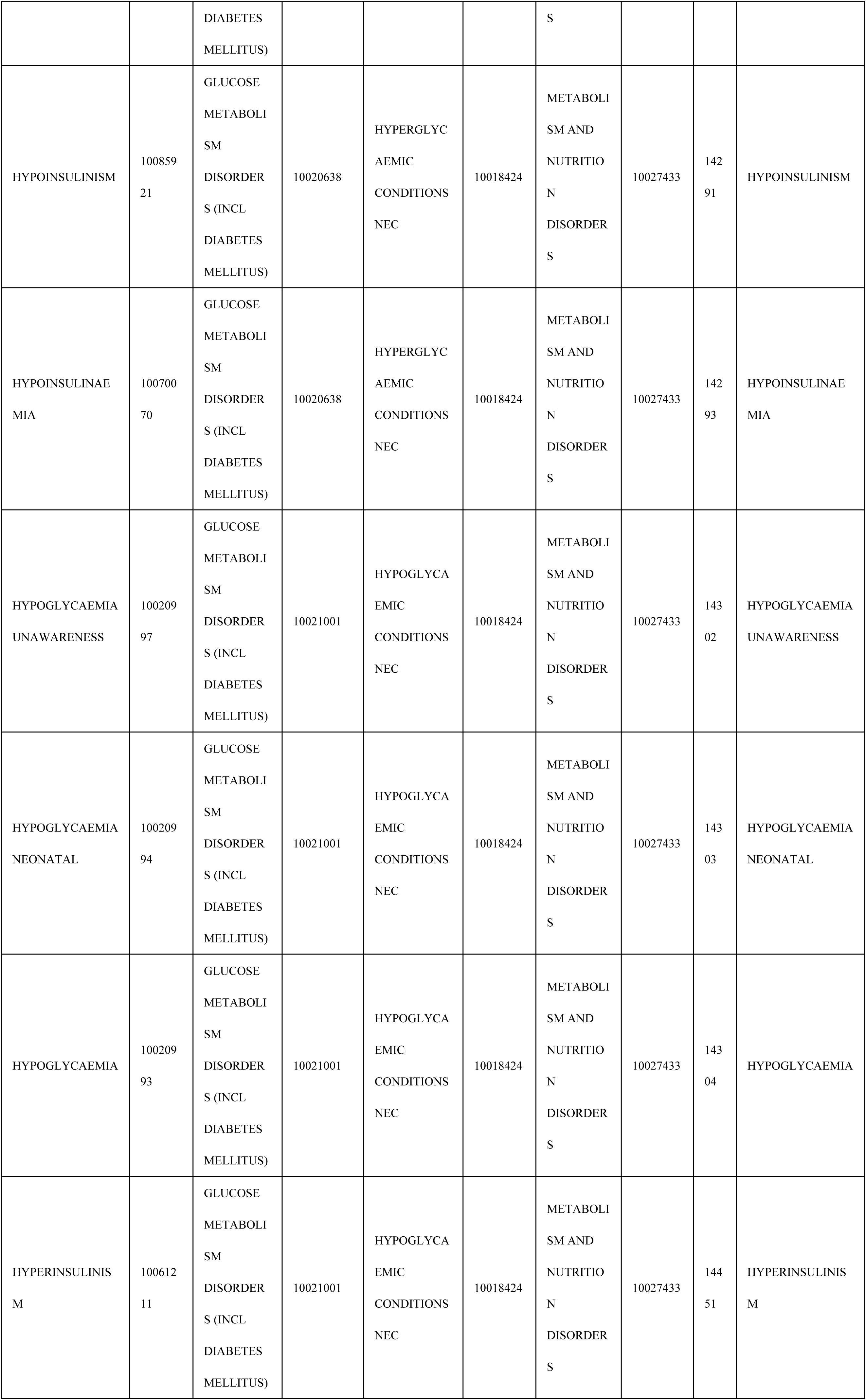
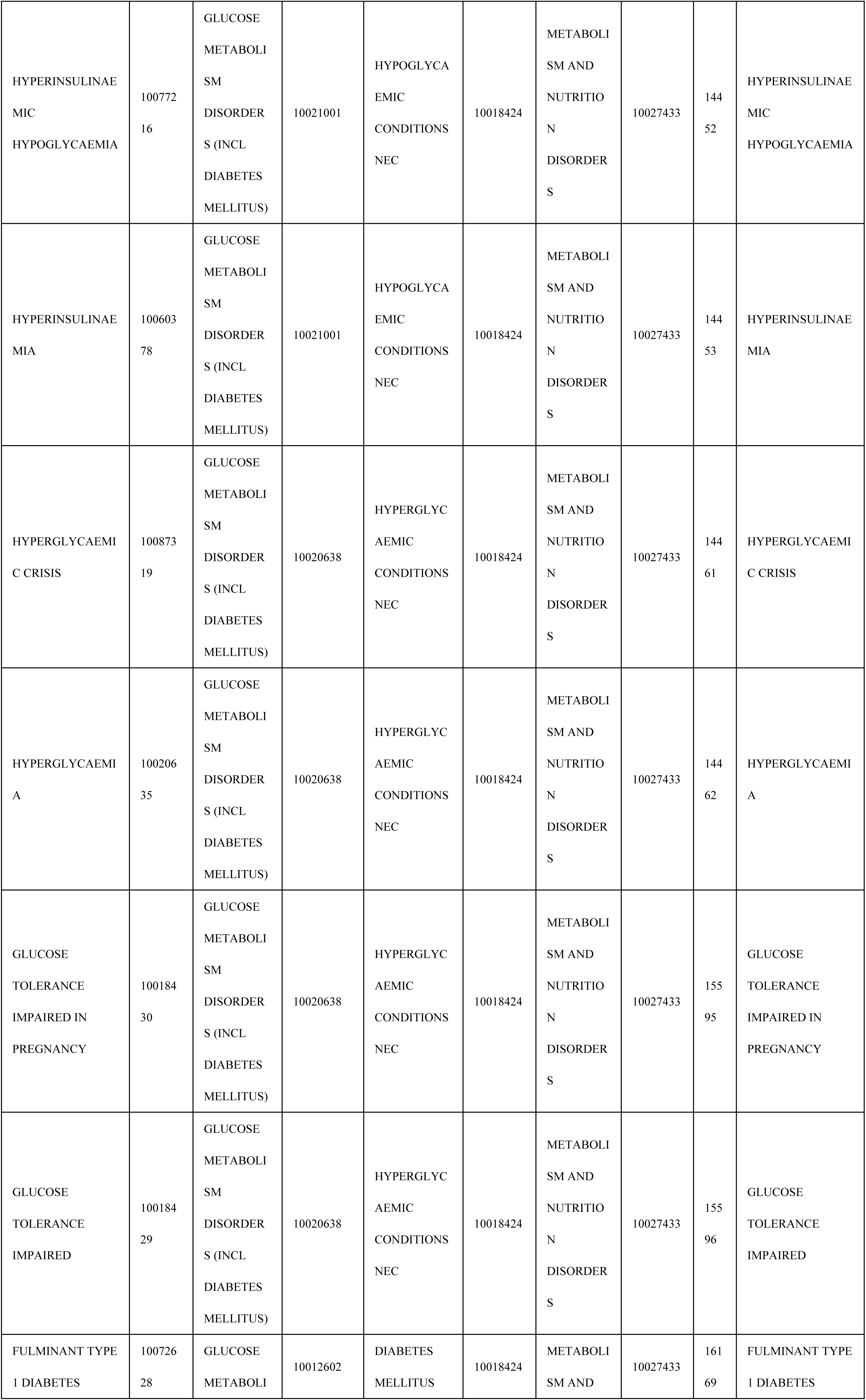
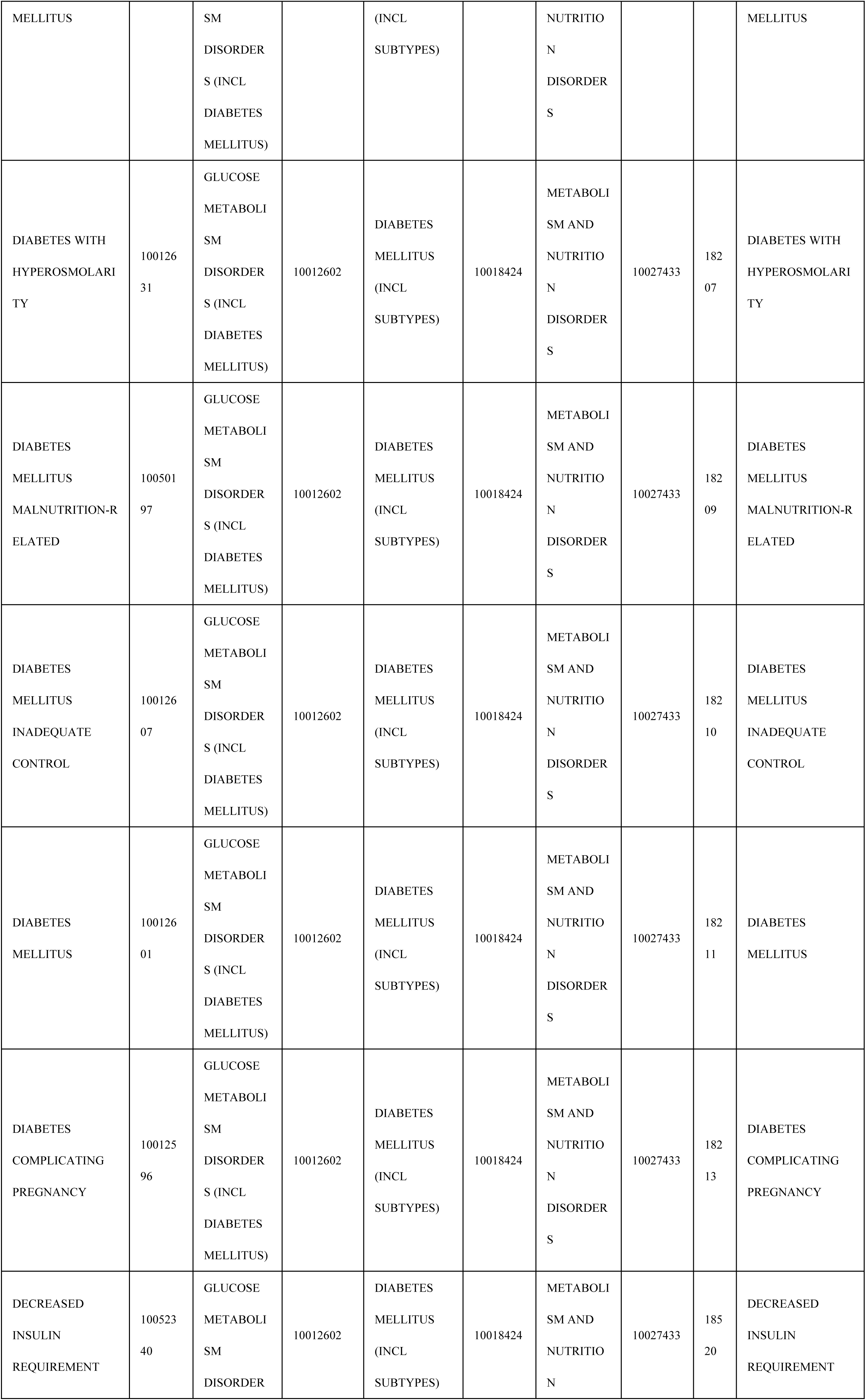
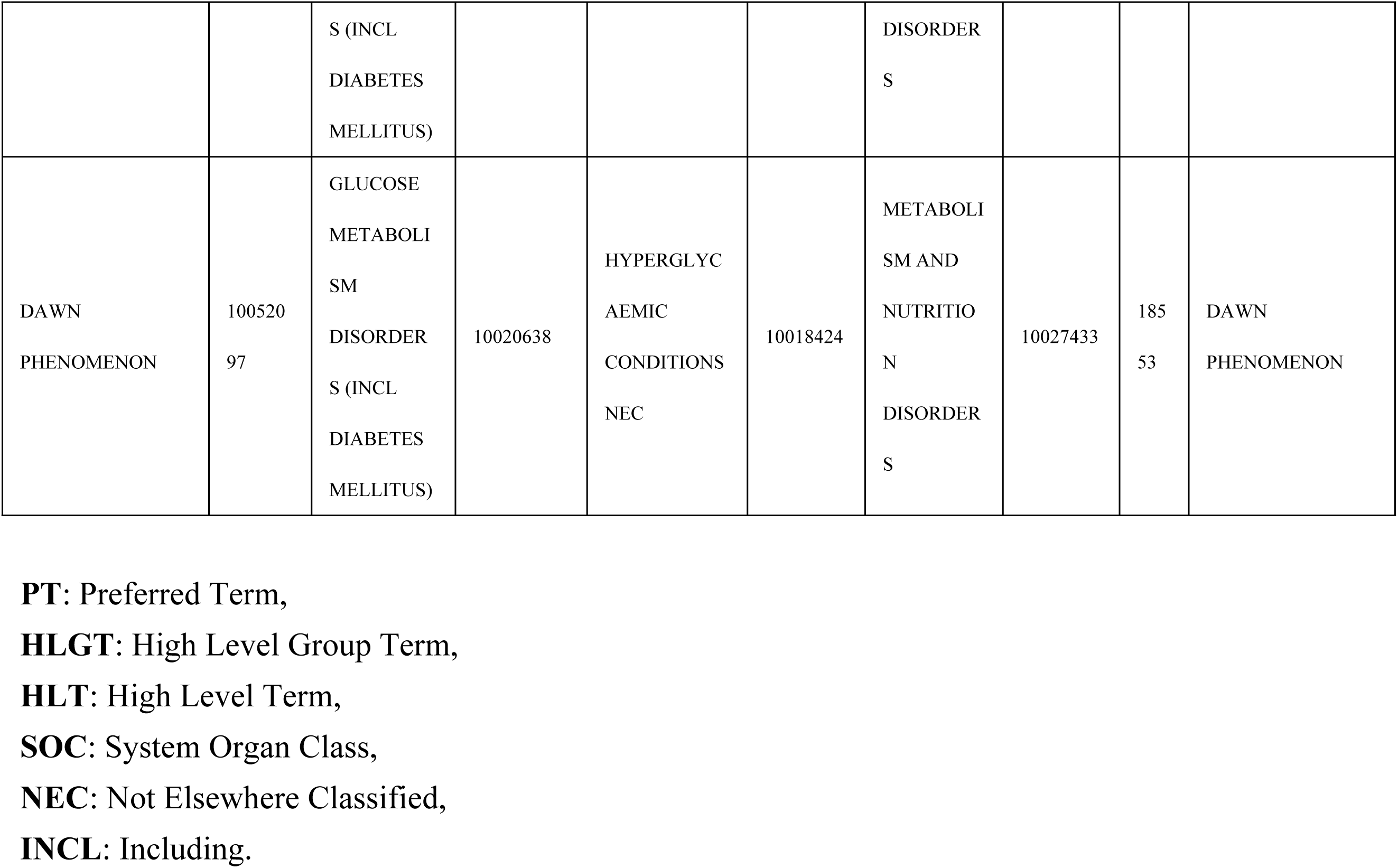
All retrieved glucose metabolism disorders contain PT signaling.

**S3 Table.**
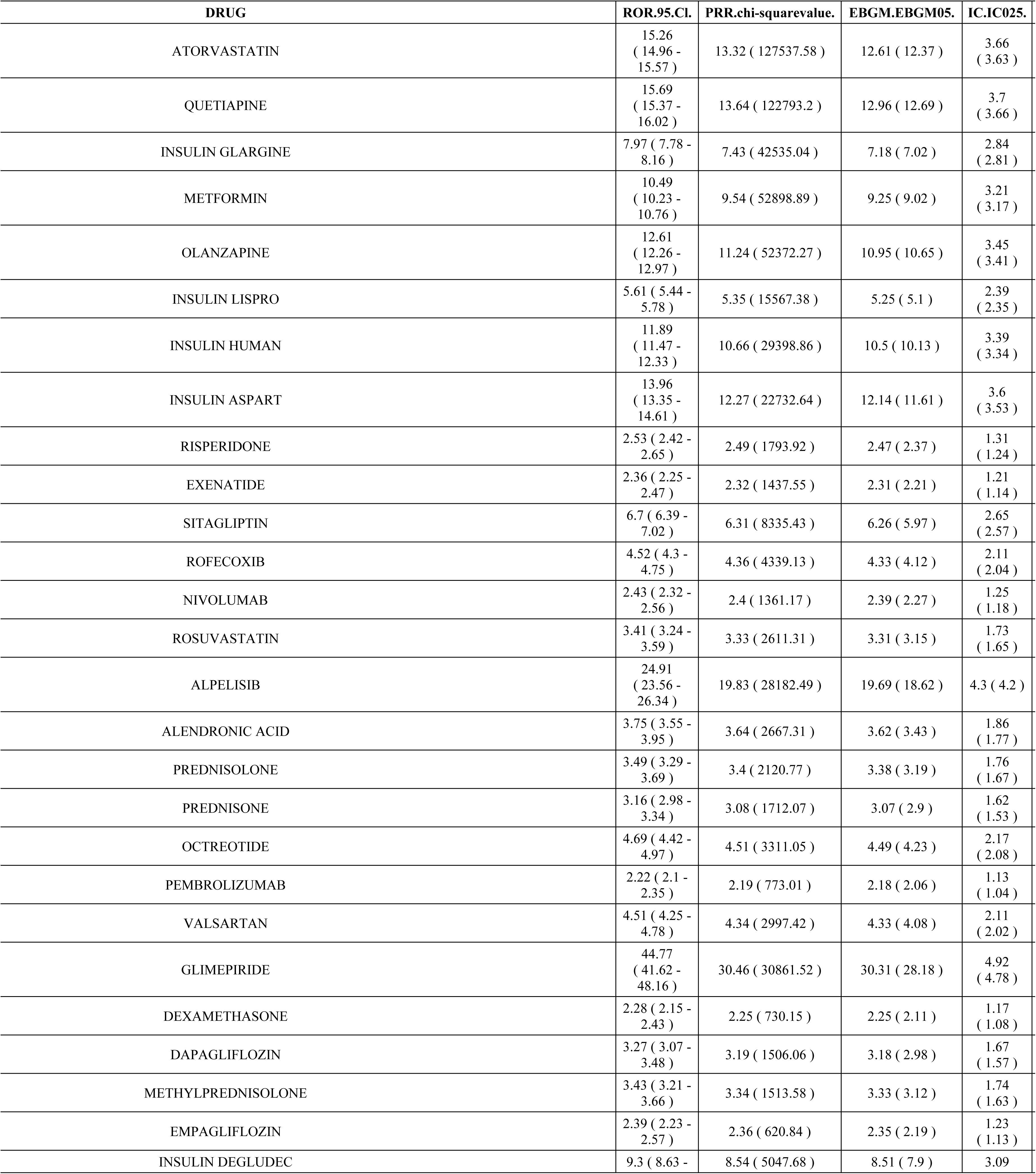
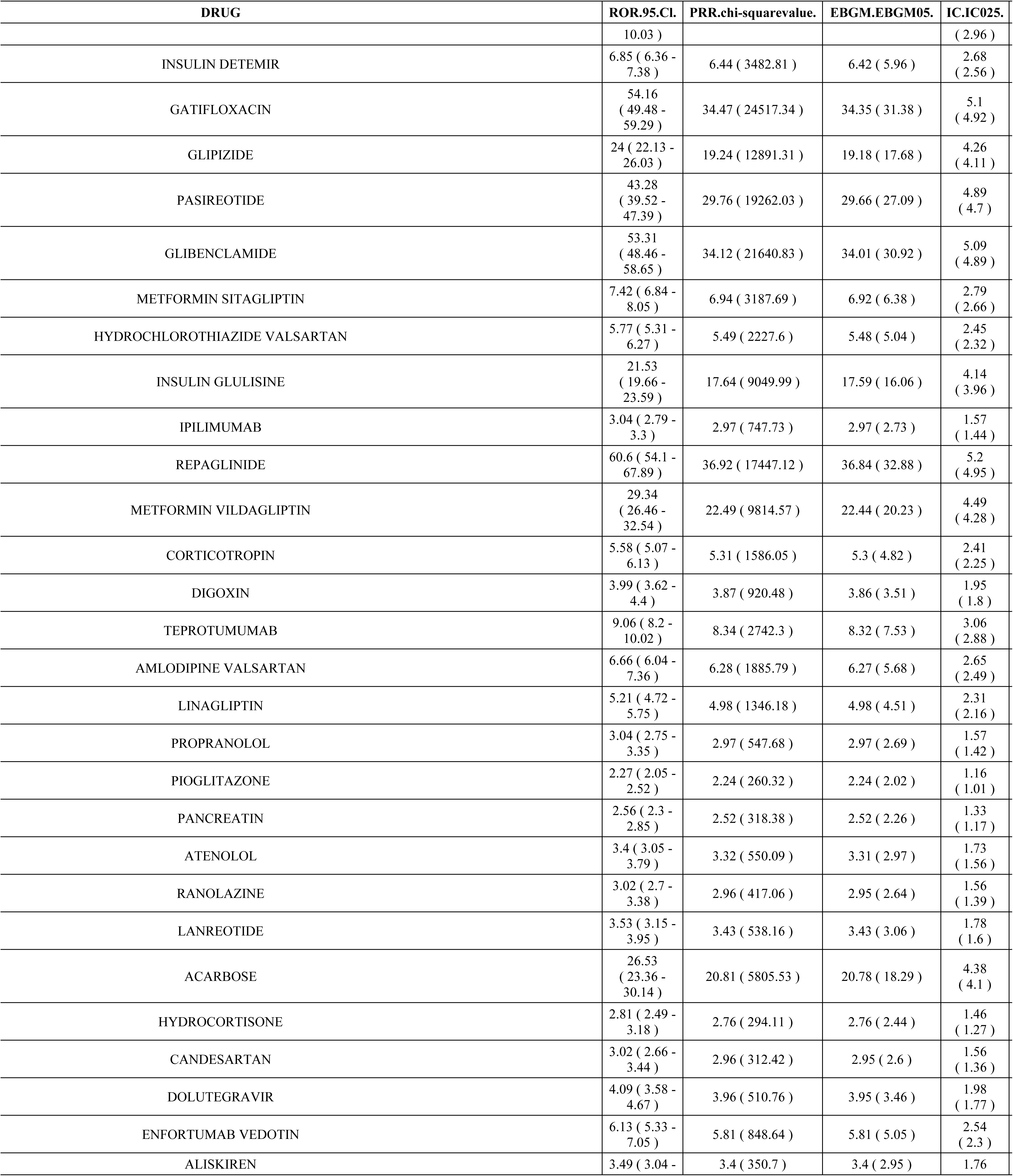
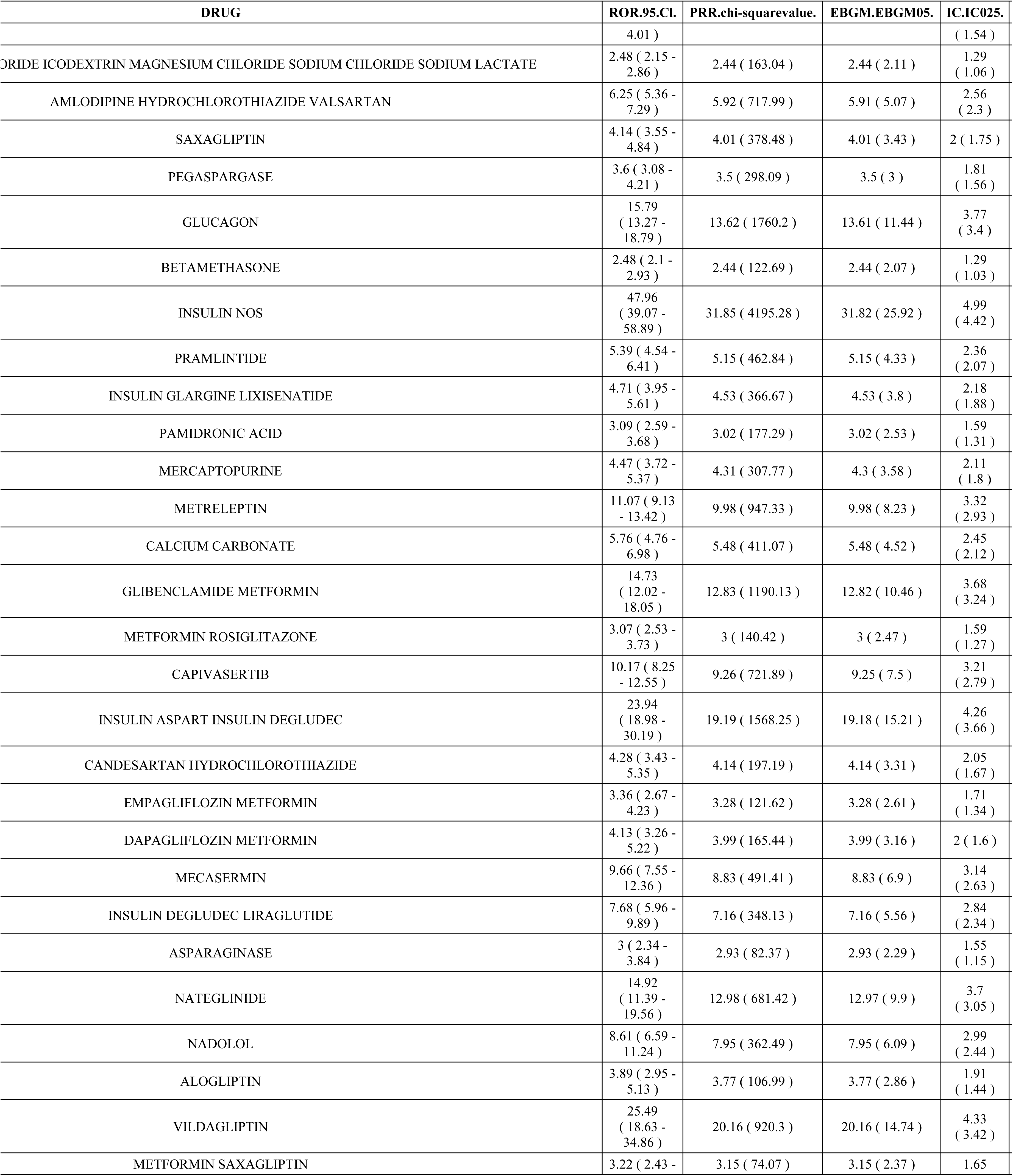
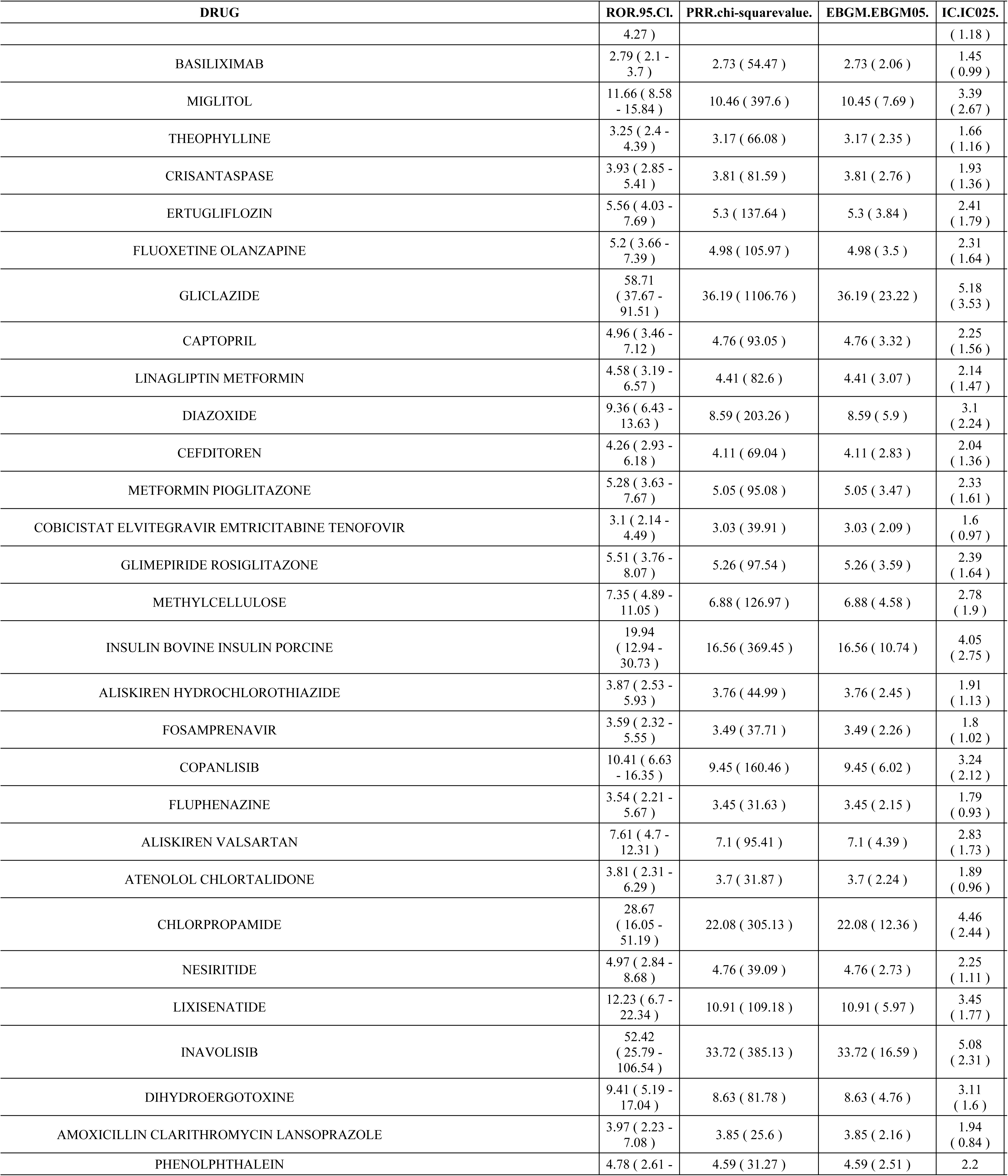
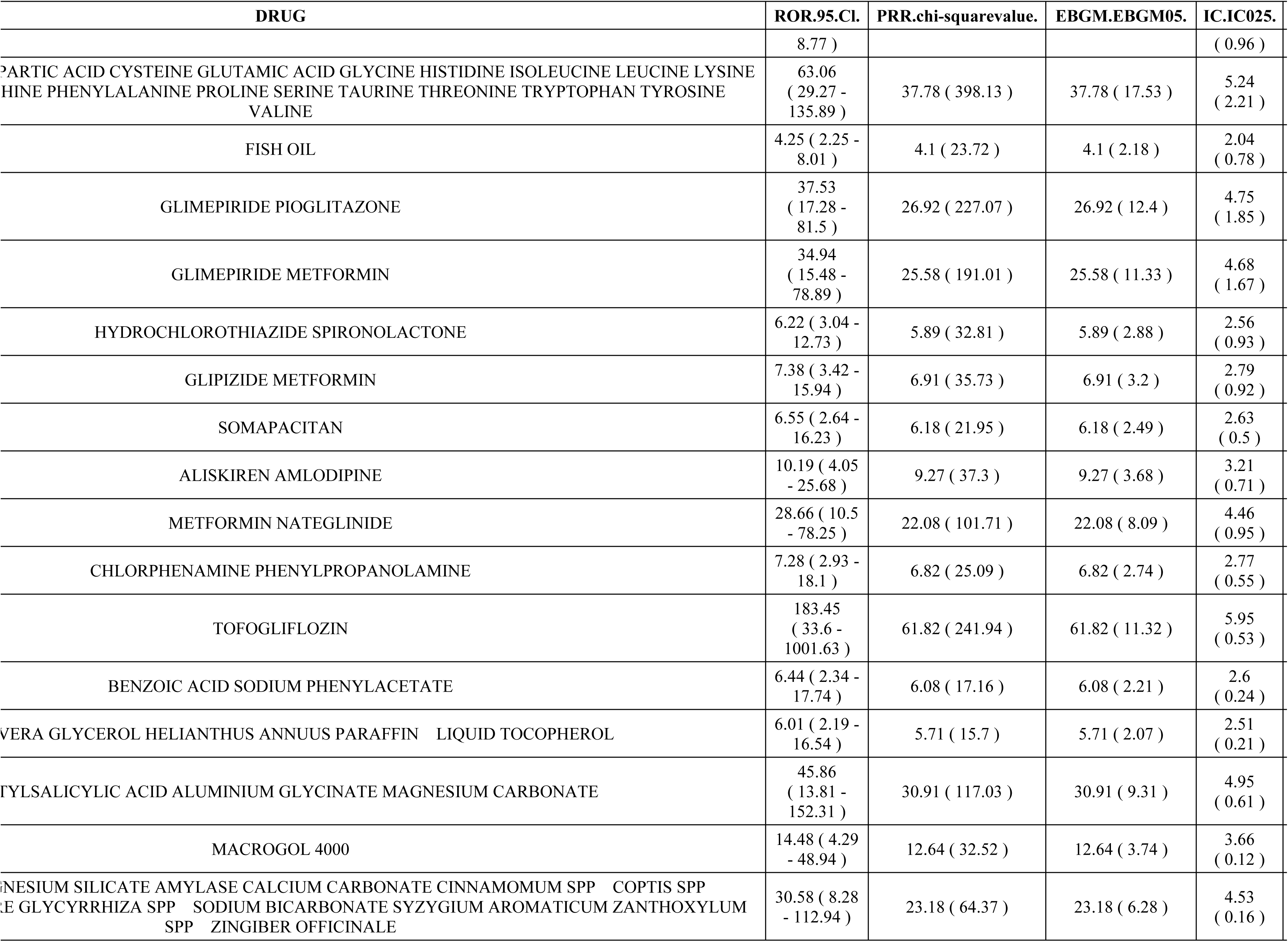
All drug results that meet the requirements of the four algorithms.

